# Genetic insights into foveal morphology and its associations with pigmentation and age-related macular degeneration

**DOI:** 10.1101/2025.06.27.25330434

**Authors:** David J. Green, Thomas H. Julian, David Romero-Bascones, Sofia Torchia, Heer N.V. Joisher, UK Biobank Eye and Vision Consortium, Unai Ayala, Maitane Barrenechea, Jay E. Self, Graeme C. Black, Tomas Fitzgerald, Ewan Birney, Constance L. Cepko, Joseph Carroll, Panagiotis I. Sergouniotis

**Author notes:** Correspondence to Panagiotis I. Sergouniotis. A list of consortium authors appears at the end of the paper.

## Abstract

The fovea is the small depression at the neurosensory retina that underlies high-resolution central vision. It is vulnerable to disease and disruption of its architecture can lead to visual disability. Foveal morphology varies substantially across individuals but the molecular causes and functional consequences of this variation remain incompletely understood. Here, we analyse six foveal morphological parameters derived from Optical Coherence Tomography (OCT) images of 39,521 UK Biobank participants. We find marked variation in foveal morphology and identify significant associations with sex and genetic ancestry. Genome-wide association studies identify 162 lead loci across the six foveal morphological parameters, implicating genes involved in pigmentation (including *TYR*, *TSPAN10* and *GPR143*) and developmental patterning (including *FGFR2*, *CYP26C1* and *CYP1A1*). Heritability estimates range from 29 to 43%. Foveal pit volume is associated with future risk of age-related macular degeneration (AMD) (hazard ratio 1.09, p = 0.026), a finding supported by Mendelian randomisation. These results establish foveal morphology as a highly heritable trait that is biologically linked to AMD.

## INTRODUCTION

The fovea is a specialized structure at the centre of the retina. It has a key role in high-acuity vision, supports most daily life activities and enables tasks such as reading and face recognition.^1,2^ The fovea is affected in a range of retinal disorders, including common adult-onset diseases such as age-related macular degeneration (AMD) (which is associated with loss of foveal cells) and paediatric conditions like albinism (which is characterized by foveal hypoplasia).^2^

The fovea has been studied for more than a century. Despite this, there are significant gaps in our knowledge, including the extent to which the morphology of the foveal pit contributes to high-acuity vision. This is partly because animal models that are routinely used in research (e.g. most murine and teleost models) lack this morphological specialization.^2^ Recent advances in genomics, data science and *in vivo* imaging technologies have transformed our ability to study complex biological structures like the fovea.^3^ Another major enabler has been the increasing availability of large-scale cohorts with a rich phenotypic scope. One example is the UK Biobank, a biomedical resource containing in-depth genetic and health information from >500,000 individuals.^4^ Many UK Biobank participants underwent enhanced phenotyping including visual function assessment (>130,000 volunteers) and imaging of the central retina (>84,000 volunteers);^5^ the latter was obtained using optical coherence tomography (OCT), a non-invasive imaging modality that rapidly generates cross-sectional retinal scans at micrometre-resolution.^6^

The most prominent morphological feature of the human fovea is the pit, corresponding to a central excavation of the inner retinal layers. Previous imaging studies have highlighted substantial morphological variability in respect to the depth, width and volume of the pit.^1,7–12^ This anatomical diversity is thought to arise from variations in foveal development. Indeed, a blunted or absent foveal pit is often seen in individuals born prematurely.^13^ Differences in foveal morphological parameters (hereafter, foveal traits) have also been reported between males and females and among individuals with different ancestral backgrounds.^7,14,15^

Previous studies have sought to investigate the genetic architecture of OCT-derived retinal traits, including central retinal thickness.^16–21^ A recent study also explored the genetic basis of foveal morphology, though it was restricted to a single parameter (pit depth).^17^ Here, we present a comprehensive, systematic analysis of six foveal traits, integrating genetic association studies, heritability estimation, genetic correlation analyses and longitudinal modelling. Together, these analyses define the genetic architecture of foveal morphology and support its relevance to common retinal disease (Fig. 1).

**Fig. 1.**
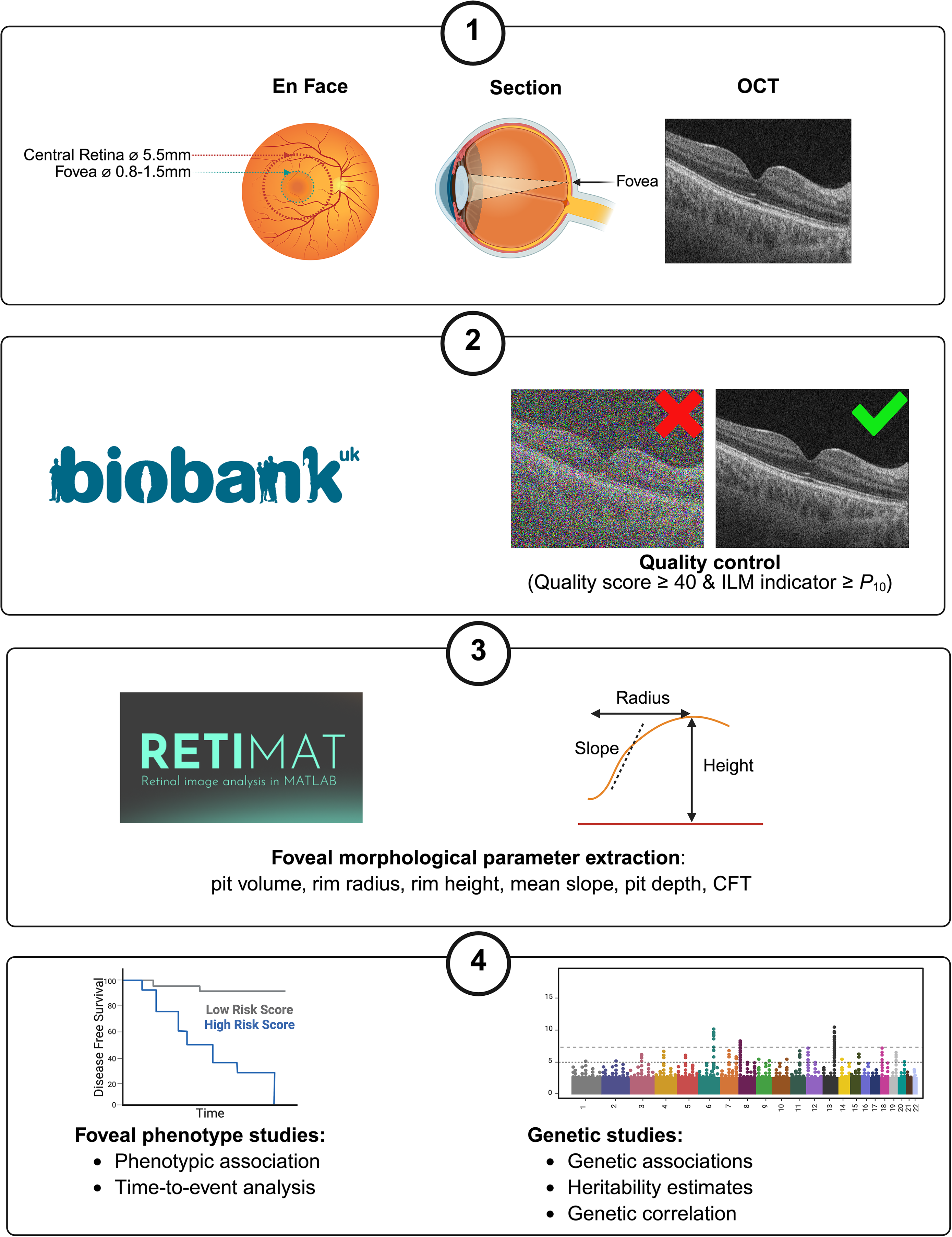
Outline of the study design. First, we used the RETIMAT tool to extract six key foveal traits from UK Biobank OCT scans (pit volume, rim radius, rim height, mean slope, pit depth and CFT). Subsequently, we performed phenotypic and genetic studies to gain insights into these foveal traits. Phenotypic analyses included the investigation of the relationships between the studied foveal traits and age, spherical equivalent refractive error, sex, genetic ancestry and visual acuity. Time-to-event analyses involved assessing the relationships between the six studied foveal traits and two major causes of visual impairment, age-related macular degeneration and glaucoma. Genetic investigations included common-variant genome-wide association studies, rare-variant burden tests, heritability estimation and genetic correlation analyses. Figure created in BioRender; Julian, T. https://biorender.com/94k714t (2026). OCT, optical coherence tomography; CFT, central foveal thickness.

## RESULTS

### Phenotypic variability in foveal traits

After applying standard OCT quality control filters^21^, we defined a subset of the UK Biobank population with high-quality OCT scans (see Methods section for further details). Restricting this subset to unrelated individuals yielded a cohort of 39,521 participants with an age and sex profile similar to that of the overall UK Biobank population.^16^ Most study subjects were female (52%) and the mean age at OCT imaging was 57.9 years (standard deviation [SD]: 8.1 years).

For each study subject, an OCT ‘volume scan’ (containing 128 cross-sectional images) was available from the central retina of each eye. These scans were processed using RETIMAT, an automated pipeline designed to extract foveal traits from OCTs.^15^ We analysed both eyes but focused primarily on the left eye as it was imaged second and showed slightly higher image quality on average.^5,19^ Six foveal parameters were assessed: foveal pit volume, rim radius, rim height, mean slope, pit depth and central foveal thickness (CFT) (Fig.1 and Table 1). The greatest fold range across UK Biobank participants was observed for pit volume (14.9-fold), followed by pit depth (6.3-fold) and mean slope (6.0-fold), indicating large differences between the most extreme phenotypes in the population. In contrast, parameters such as rim height (1.3-fold), rim radius (2.1-fold) and CFT (3.0-fold) were more constrained (Fig.2).

**Fig. 2.**
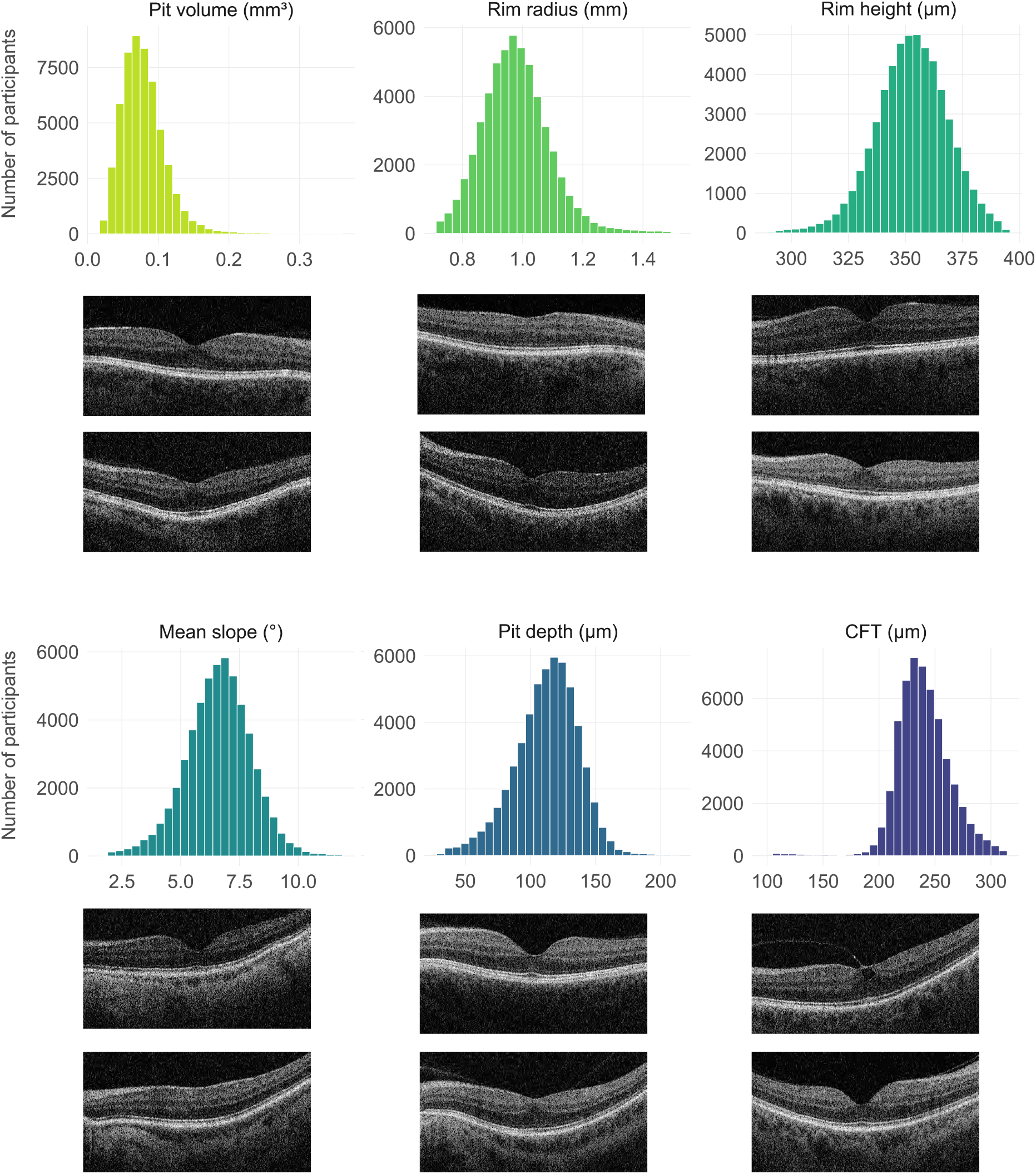
Histograms showing the distribution of six foveal traits in UK Biobank participants (left eyes; after removal of 1% from the upper and lower tails of the data). Also shown are six examples of OCT scans from the extreme ends of each distribution (reproduced with the permission of UK Biobank). The following mean values were obtained: pit volume 0.079 mm^3^ (range: 0.024–0.352 mm^3^), rim radius 0.98 mm (range: 0.71–1.48), rim height 353 µm (range: 291–394 µm), mean slope 6.62° (range: 1.93–11.6°), pit depth 113 µm (range: 33.4–209 µm), central foveal thickness (CFT) 240 µm (range: 104–312 µm). A summary of the mean and median values for all traits are provided in Supplementary Tables 2 and 3. The data underlying the histograms, including the number of participants in each bin, are provided in the Supplementary Data file. The y-axis indicates the number of participants in each bin.

**Table 1.**
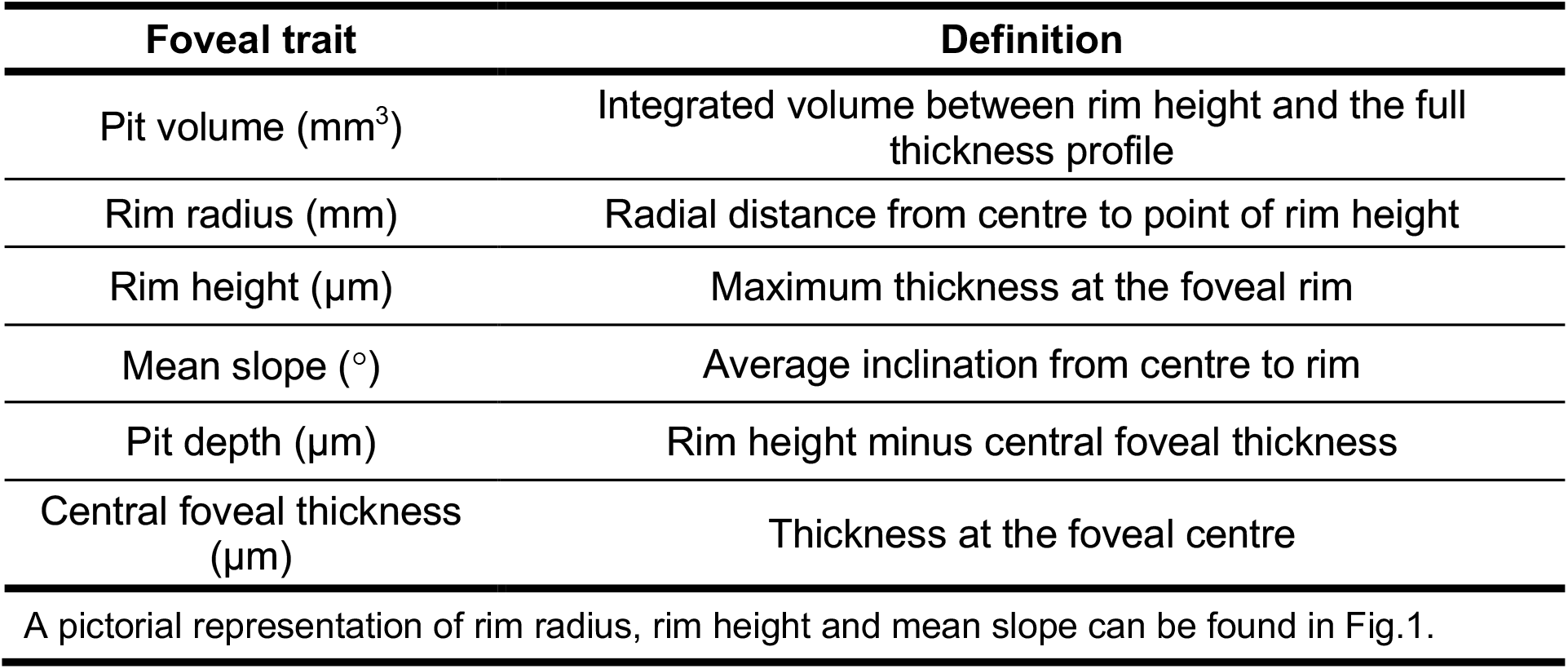
Definitions of the six RETIMAT-derived foveal traits analysed in this study.

We next used multivariable linear regression models to assess how key demographic and ophthalmic factors relate to foveal morphology (Fig.3). Across traits, spherical equivalent refractive error showed modest associations and generally small effect sizes. In contrast, sex had a clear and consistent influence on foveal traits: female sex was associated with greater pit volume, rim radius and pit depth. Genetic ancestry also accounted for substantial variation; individuals of African ancestries differed from their European counterparts in several traits and, on average, had larger pit volume, rim radius and pit depth.

**Fig. 3.**
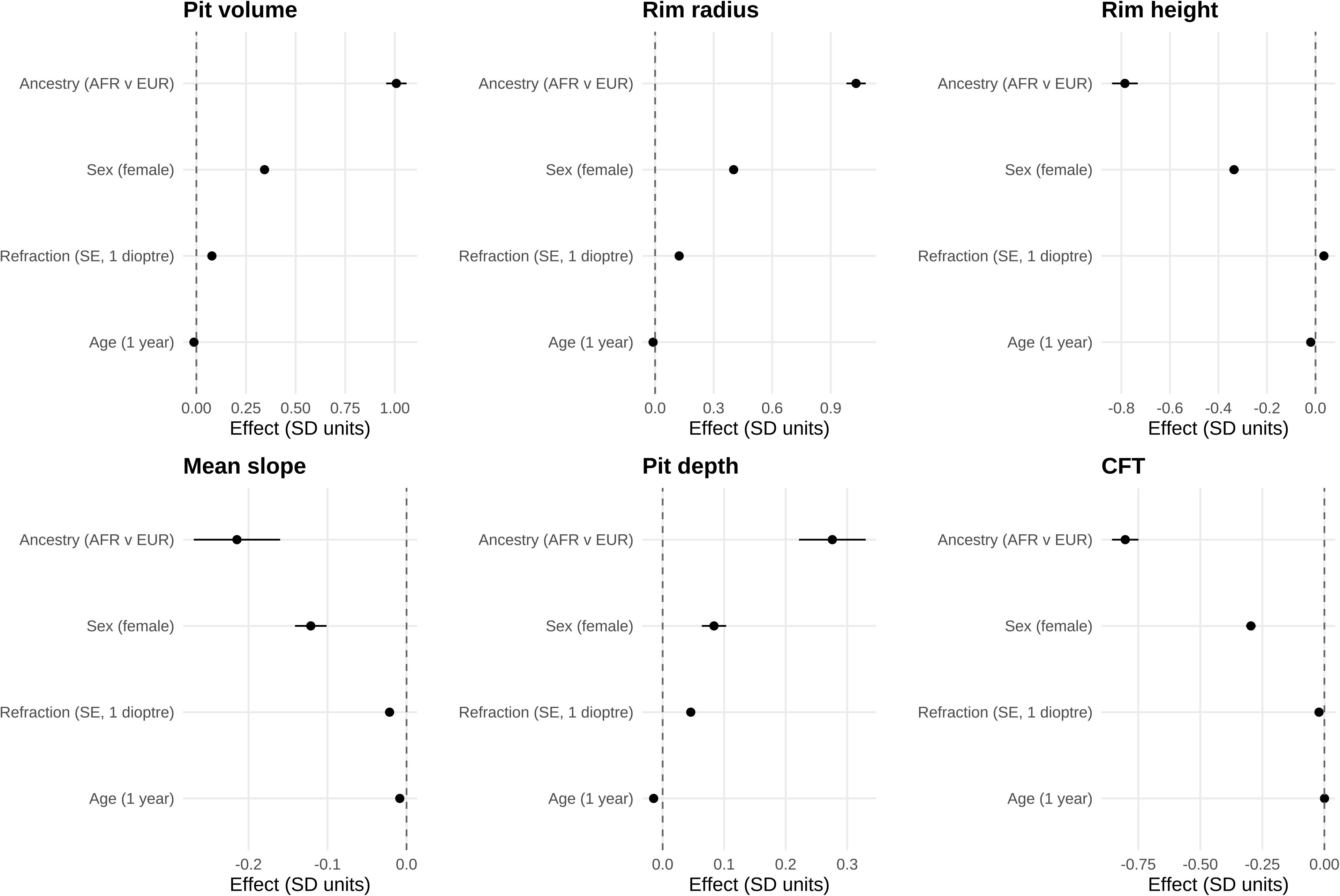
Forest plots showing the results of multivariate linear regression analyses for foveal traits in relation to key participant characteristics (genetic ancestry, sex, refractive error and age) (n=39,521). Each panel shows the effect estimates from linear regression models fitted separately for the six foveal traits. Points represent regression coefficients, with horizontal bars indicating 95% confidence intervals. Sex and ancestry showed the largest and most consistent effects across traits, while age and refractive error exhibited comparatively smaller influences. Full numerical results are provided in Supplementary Table 1. While this analysis used left-eye scans, averaging measurements from both eyes produced comparable results (Supplementary Fig. 1).

Taken together, these results highlight sex and ancestry as key drivers of inter-individual variability in foveal morphology, with spherical equivalent and age having more modest effects (Supplementary Tables 1-3).

Aiming to gain insights into the factors underlying the detected differences between ancestral groups, we investigated the role of fundus (principally choroidal) pigmentation. To evaluate this, we used a fundus imaging-derived proxy of pigmentation, the ‘Retinal Pigment Score’ (RPS).^22^ We then compared the change in adjusted R² (a measure of model fit) associated with adding RPS or ancestry across the six studied foveal traits; age, sex and spherical equivalent refractive error were included as covariates. Overall, addition of genetic ancestry was associated with larger improvements in model fit than addition of fundus pigmentation for most traits, particularly pit volume and rim radius. It is noted that addition of pigmentation produced little change in adjusted model fit for rim radius, suggesting limited additional explanatory value beyond genetic ancestry (Fig.4; Supplementary Table 4).

**Fig. 4.**
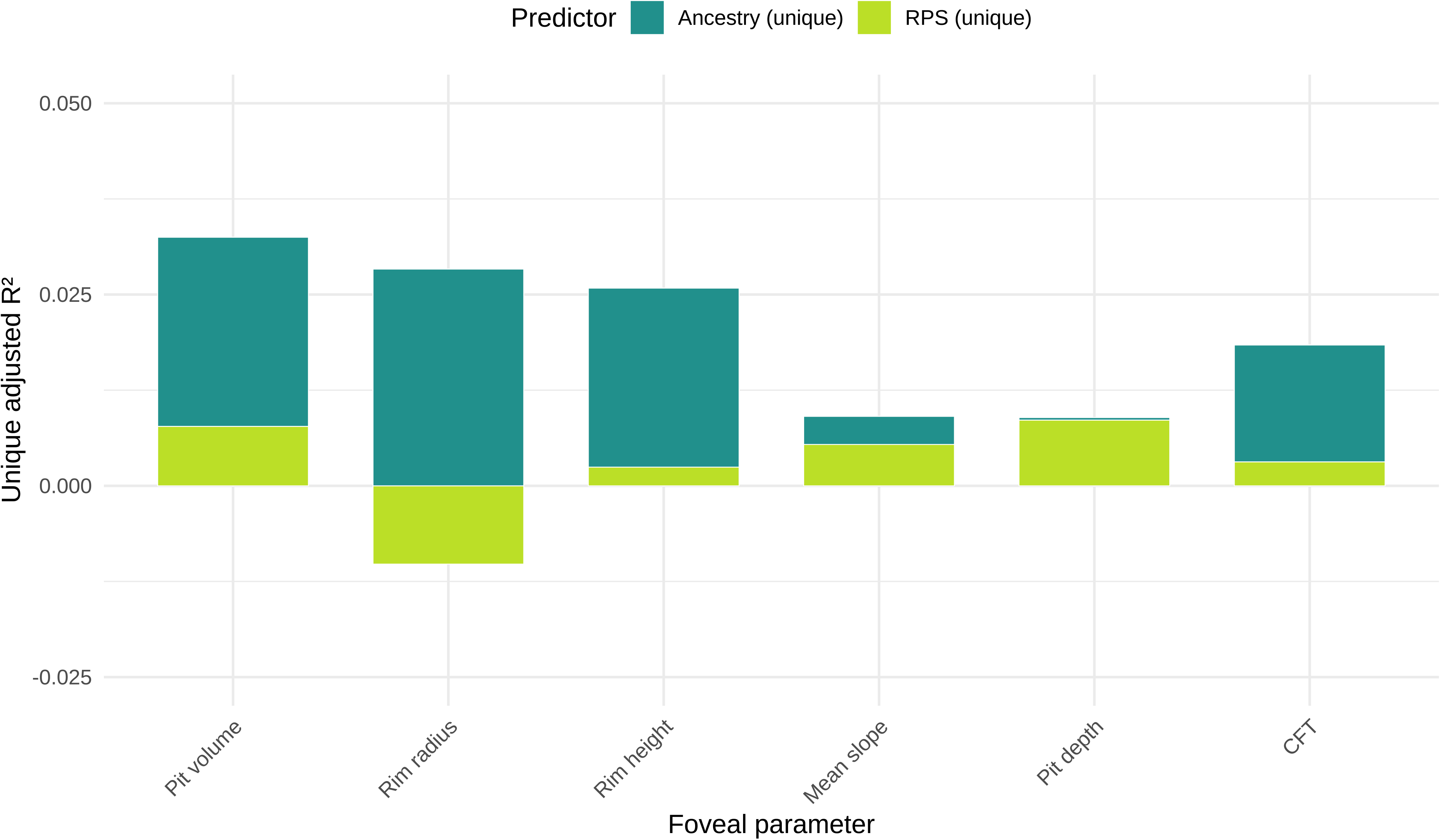
Relative influence of fundus pigmentation and genetic ancestry on foveal morphology (left eye). Bar plots show the change in adjusted R² values obtained by adding Retinal Pigment Score (RPS) or genetic ancestry to linear models of the six foveal traits (after adjustment for age, sex and spherical equivalent refractive error). Values were calculated by comparing nested models containing RPS, ancestry, or both variables. Across most traits, addition of genetic ancestry was associated with larger increases in adjusted model fit than addition of RPS. The small negative value observed for rim radius indicates that inclusion of RPS did not improve adjusted model fit in that model. Full numerical results are provided in Supplementary Table 4.

To assess the extent to which foveal morphology predicts visual acuity, we performed linear regression with LogMAR vision as the outcome, and age, sex, spherical equivalent refractive error and genetic ancestry as covariates. All six foveal traits showed statistically significant associations with acuity except central foveal thickness, but in every case the effect sizes were small (model R² values ranged from 2.8 to 3.0%). Mean slope was the strongest predictor, with flatter slopes associated with worse vision. Full effect estimates and confidence intervals are provided in Supplementary Fig. 2.

### Association between foveal traits and risk of retinal disease

To investigate whether foveal morphology constitutes a risk factor for future retinal conditions, we performed Cox proportional hazards modelling using the six studied foveal traits, along with time-to-event data for two leading causes of visual impairment, AMD and glaucoma. We adjusted for age, sex, spherical equivalent refractive error and genetic ancestry in these analyses.

For AMD (n=959 cases), significant associations were observed with larger pit volume (hazard ratio [HR] per one SD 1.09; 95% confidence interval [CI] 1.04– 1.16; p = 0.026). For glaucoma (n=844 cases), there were statistically significant protective effects for mean slope (HR per SD 0.90; 95% CI 0.84–0.96), rim height (HR per SD 0.83; 95% CI 0.77–0.89) and pit depth (HR per SD 0.87; 95% CI 0.82–0.93) (Fig.5, Supplementary Table 5).

**Fig. 5.**
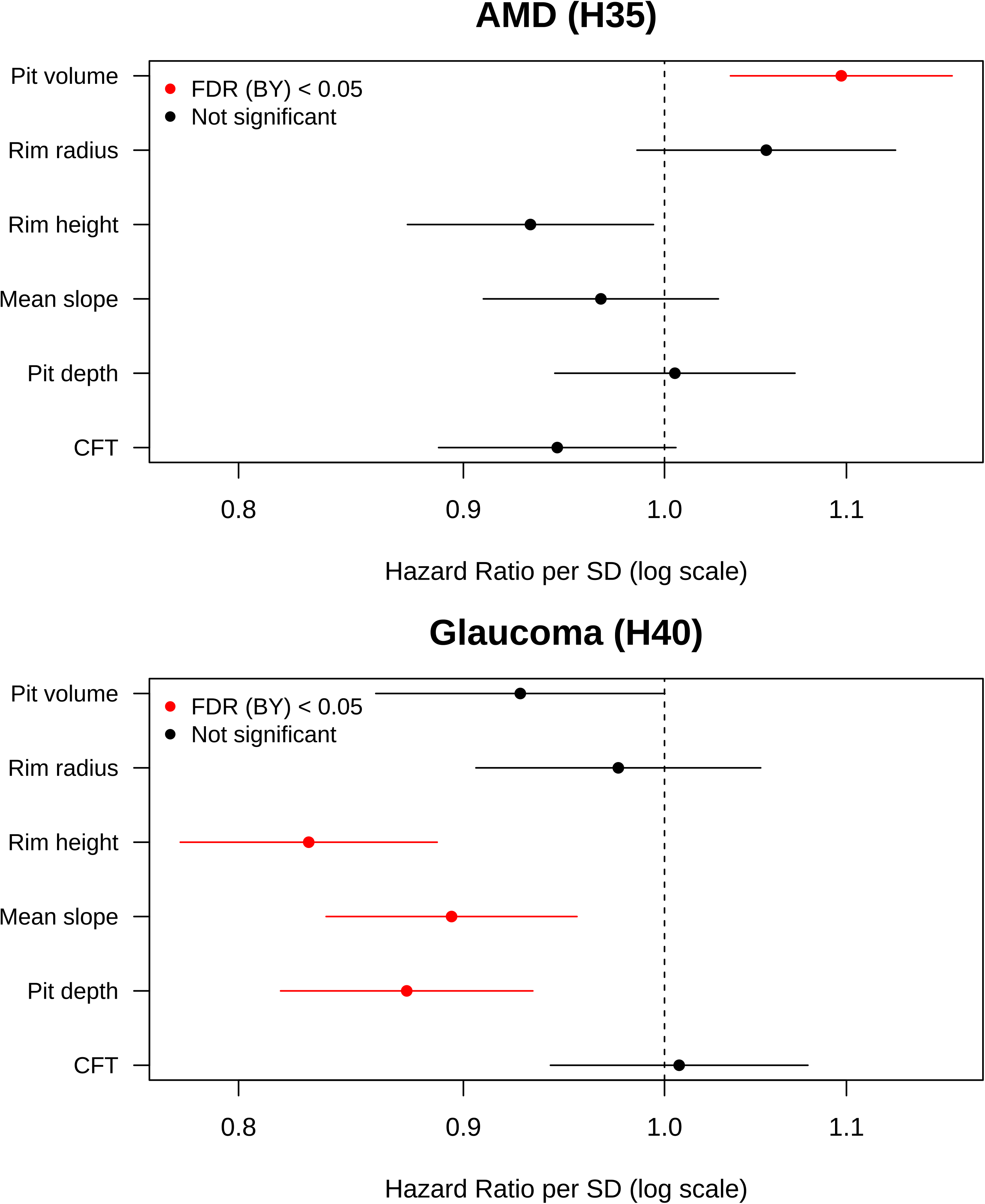
Cox regression analysis of the associations between foveal traits and post-OCT-scan assigned codes for AMD (H35) and glaucoma (H40) (n=39,521). Associations meeting the 5% false discovery rate Benjamini-Yekutieli threshold (two-sided tests) are highlighted in red. Further information, including numerical data, can be found in Supplementary Table 5. While this analysis used left-eye scans, averaging measurements from both eyes produced comparable results (Supplementary Fig. 3). The foveal traits studied are closely related (Fig.7), so their independent contributions are difficult to disentangle. The association observed for pit volume may therefore reflect a broader correlated axis of foveal morphology (rather than a specific causal role of pit volume). AMD, age-related macular degeneration; CFT, central foveal thickness; SD, standard deviation; OCT, optical coherence tomography.

### Genetic associations of foveal traits

To investigate the genetic factors influencing the six studied foveal traits, we performed common-variant genome-wide association studies (GWAS) using REGENIE.^23^ For these analyses, we defined a subset of the UK Biobank population that can be considered genetically well-mixed (*i.e.* includes participants that were assigned by genotype principal component analysis to a cluster with subjects of mostly European-like ancestries). After applying relevant imaging, genetic and foveal trait quality controls, this cohort included 36,205 individuals; 29,710 of these were recruited between 2006 and 2010 (Instance 0, ‘Initial assessment’) and were used in the primary analysis while 6,495 were independently recruited between 2012 and 2013 (Instance 1, ‘First repeat assessment visit’) and were included in the internal validation study. The following covariates were incorporated into the models: age, sex, height, weight, spherical equivalent refractive error and genetic principal components 1 to 20. A study-wide threshold (p < 8.3 × 10^−9^) was used to account for the different association routes that were utilized; this was obtained after applying Bonferroni correction for six tests to the conventional p < 5 × 10^−8^ ‘genome-wide significance’ threshold.

Genome-wide significant variants in the primary GWAS included: 5,023 for pit volume; 5,234 for rim radius; 6,730 for rim height; 3,457 for mean slope; 4,006 for pit depth; and 2,387 for CFT. Following analysis with GCTA-COJO (conditional and joint multiple-variant analysis),^24^ these merged into the following lead loci: 54 for pit volume, 56 for rim radius, 35 for rim height, 27 for mean slope, 35 for pit depth and 23 for CFT (Fig.6, Supplementary Fig.4-9; Supplementary Data). We subsequently performed an internal validation GWAS with this showing a high level of concordance with the primary analysis (all traits had a correlation coefficient greater than 0.9; Supplementary Fig.10–15). Although the internal validation sample was considerably smaller, several lead loci still reached genome-wide significance in both studies: 6 for pit volume, 3 for rim radius, 1 for rim height, 1 for mean slope and 1 for pit depth.

**Fig. 6.**
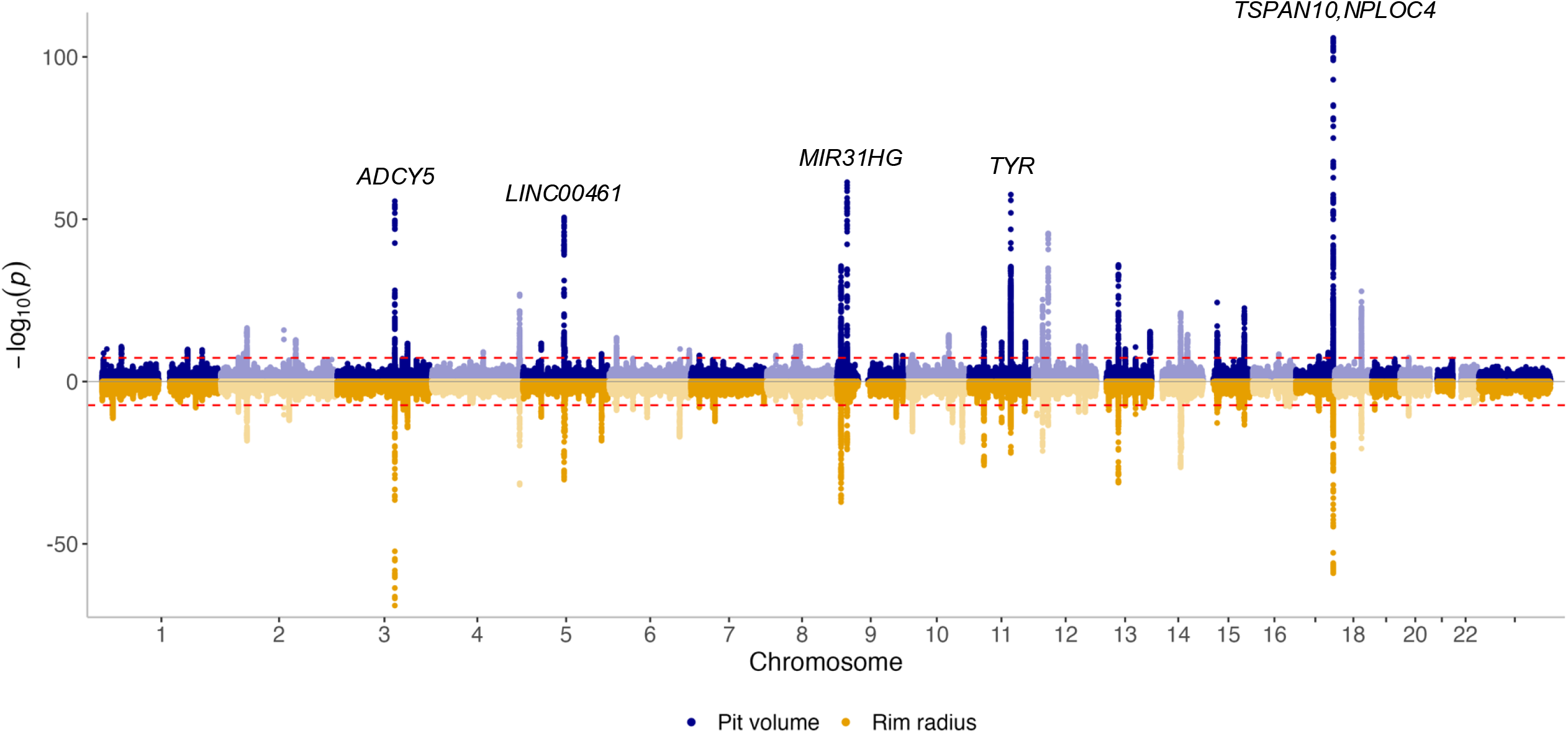
A Miami plot, illustrating the results of the primary genome-wide association study for foveal pit volume (top) and rim radius (bottom) (left eye). The red line indicates the threshold for genome-wide significance and 5 key loci exceeding this value have been annotated.

We identified a combination of trait-specific and shared loci influencing foveal morphology. Many of these loci encompass variants previously linked to retinal layer thickness parameters (including *TSPAN10* and *LINC00461*) while a subset of them has been linked to monogenic disorders, most notably, albinism (including *TYR*, *OCA2* and *GPR143*) (Table 2 and Supplementary Data).

**Table 2.**
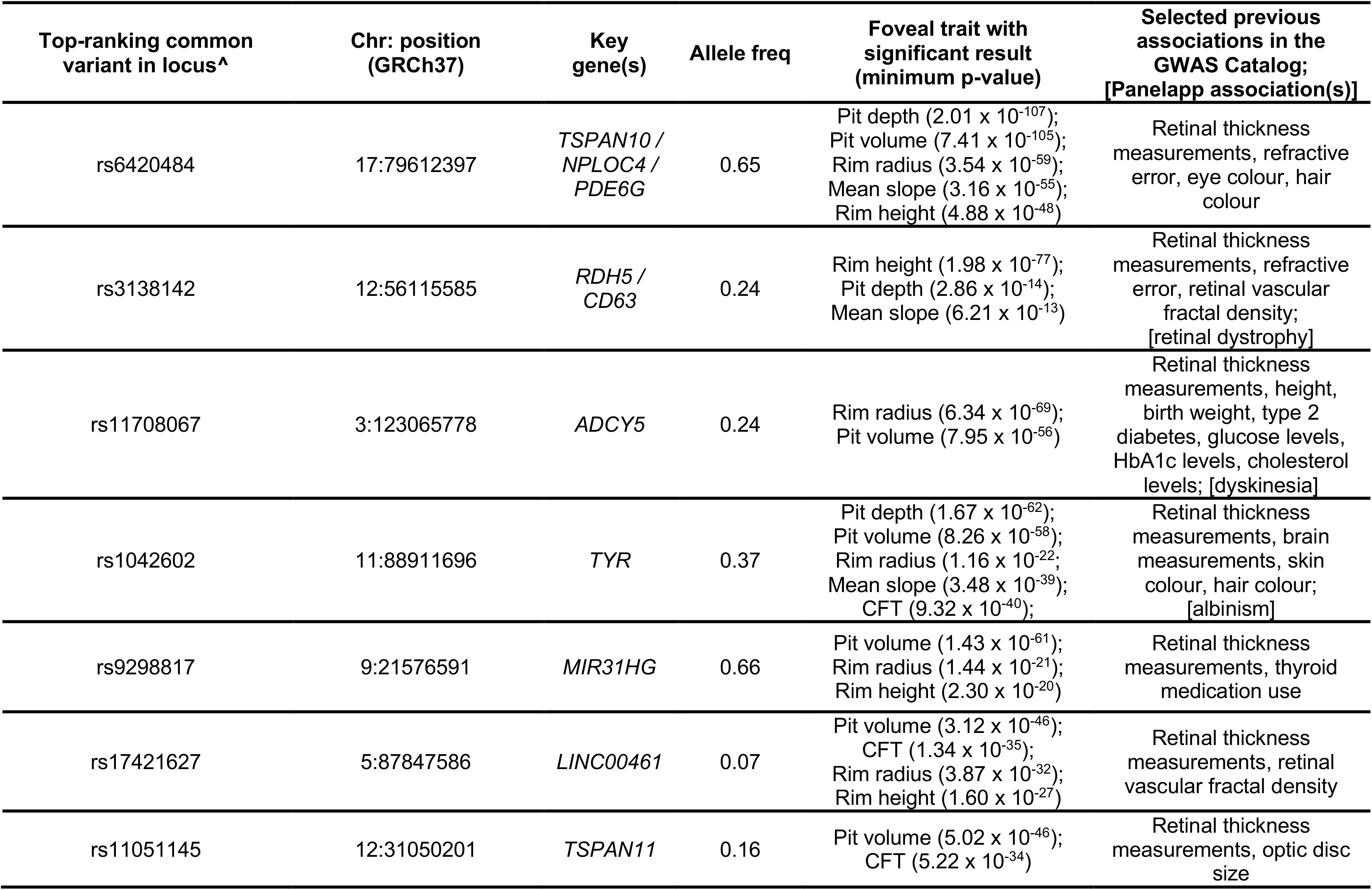
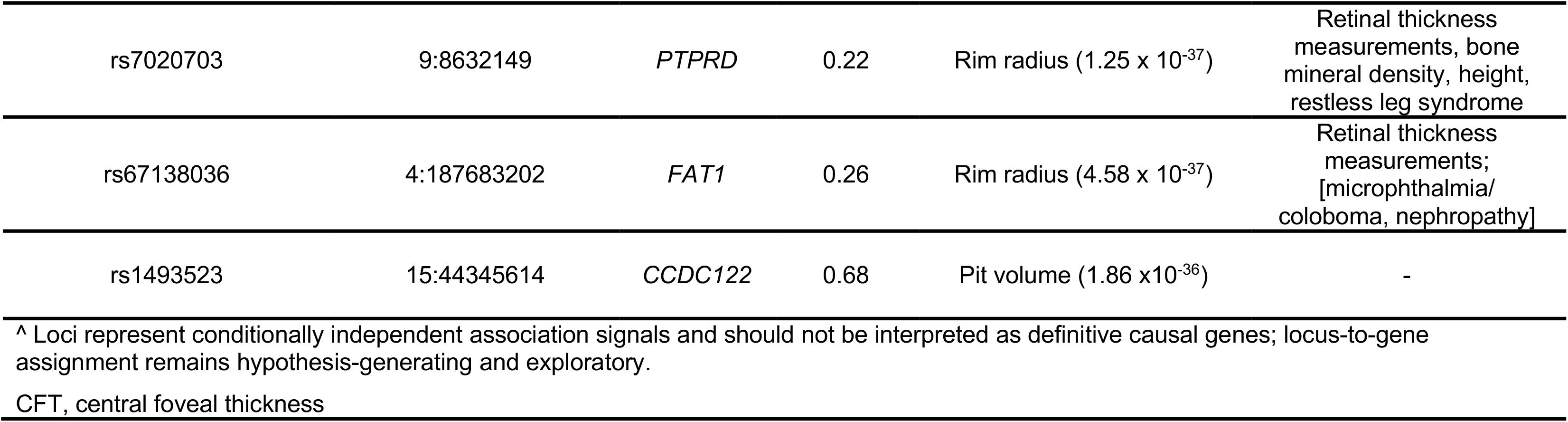
Summary of the 10 top-ranking loci associated with foveal traits.

Furthermore, we identified several lead signals that have not been previously linked with retinal phenotypes (including variants at the *CYP1A1* and *KMT2A* loci). We also detected multiple lead variants for which associations have only been detected for retinal thickness measurements but not for foveal/macular traits (including at the *FGFR2* and *SPRY2* loci).

MAGMA gene set enrichment analysis did not yield any statistically significant hits after correction for multiple testing. We note that the genomic inflation factors (λGC) ranged from 1.12 to 1.17, with corresponding linkage disequilibrium (LD) score intercepts close to 1 (range: 1.005–1.027). These results suggest minimal confounding from population structure or cryptic relatedness. The attenuation ratios were uniformly low (all < 0.11), consistent with polygenic architecture and well-controlled inflation across all six studied traits (Supplementary Table 6).

In addition to the above common-variant GWAS, we performed a rare variant burden analysis using REGENIE.^23^ This identified genome-wide significant associations between *SIX6* and both pit depth and pit volume (p < 4.6 × 10⁻⁷).

### Heritability estimates for foveal traits

We used linkage disequilibrium (LD) score regression analysis^25^ to obtain heritability estimates for the six studied foveal traits. We found moderate to high heritability, ranging from h² = 0.29 (standard error [SE] = 0.03) for CFT to h² = 0.43 (SE = 0.04) for pit volume. Rim height (h² = 0.41), rim radius (h² = 0.39), pit depth (h² = 0.34) and mean slope (h² = 0.31) were also significantly heritable (Supplementary Table 6).

### Global genetic correlation between foveal traits

LD score regression was also used to assess the global genetic correlation between the studied traits. Because the six foveal traits capture related aspects of macular architecture, we expected some degree of phenotypic correlation. We therefore used LD score regression to quantify the extent of shared genetic architecture and to determine whether relationships between traits simply reflected measurement interdependence or suggested deeper biological overlap. We found significant interdependence between many foveal traits. The strongest correlations (rg > 0.8) were observed between pit depth and mean slope (rg = 0.88) and between pit volume and rim radius (rg = 0.85), suggesting that these pairs reflect closely related biological processes. Moderate correlations (rg = 0.6-0.8) included pit depth and pit volume (rg = 0.76), mean slope and rim height (rg = 0.65), and pit depth and rim height (rg = 0.57). Together, these indicate shared underlying variation across several morphological aspects of the fovea.

In contrast, correlations between CFT and other parameters were more variable. We observed strong negative correlations between CFT and pit volume (rg = – 0.79), pit depth (rg = –0.69) and rim radius (rg = –0.51), consistent with an inverse relationship between retinal thickness and pronounced foveal pit excavation (Fig.7).

**Fig. 7.**
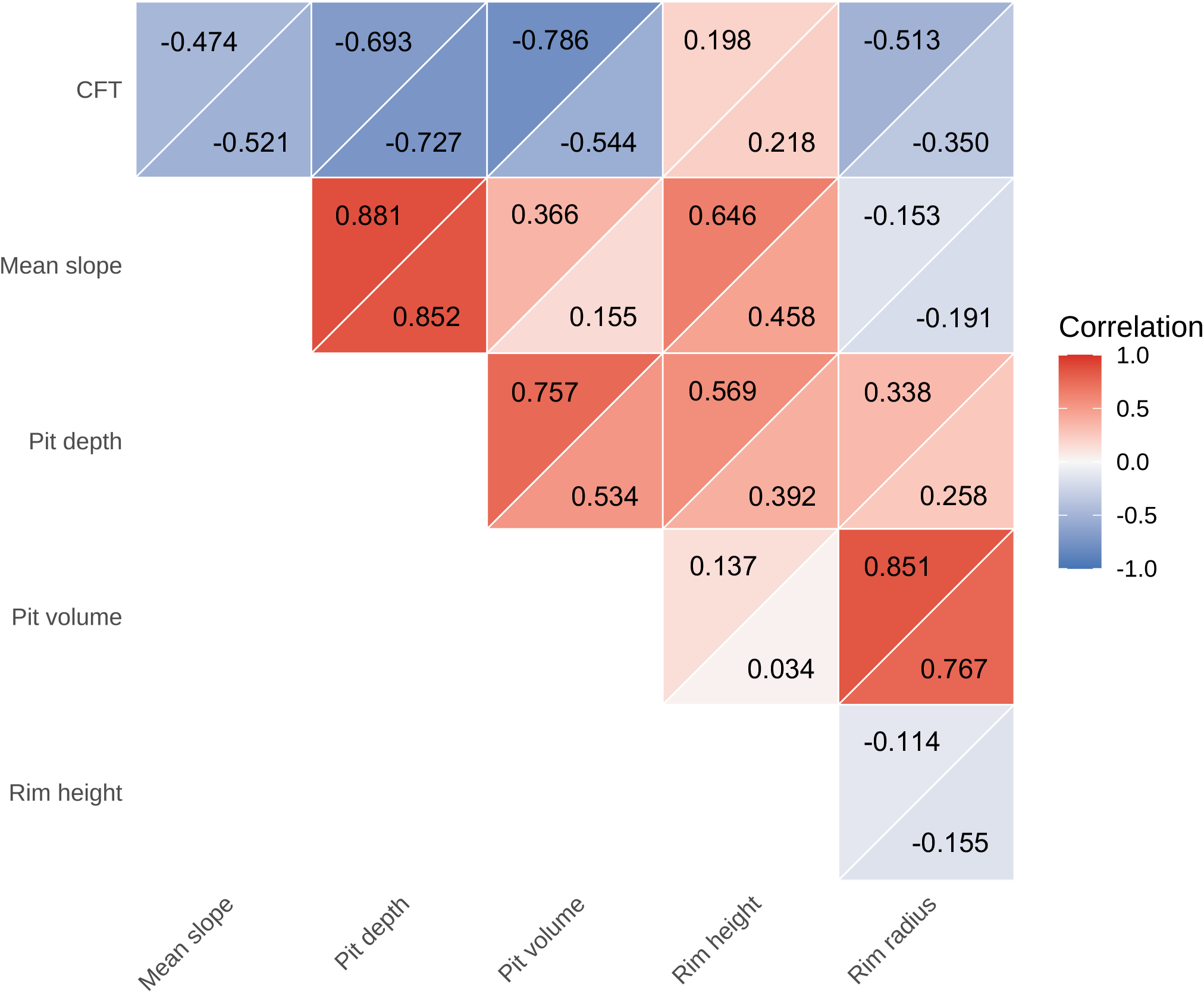
Genetic and phenotypic correlations between foveal traits. The upper triangle of each cell contains the phenotypic correlation coefficient for each trait-trait pair, whereas the lower triangle contains the corresponding genetic correlation coefficient. The observed correlation patterns are consistent with expected geometric relationships and shared genetic influences (rather than arising solely from measurement covariance). Further information, including numerical data, can be found in Supplementary Table 7. CFT, central foveal thickness.

Correlations between foveal traits and fundus pigmentation (RPS) were generally weak (rg = -0.23 with pit depth, -0.22 with mean slope), with the strongest positive correlation seen with CFT (rg = 0.23) (Supplementary Table 7).

### Mendelian randomisation

We performed Mendelian randomisation (MR) analysis on the foveal traits that we found to be associated with AMD or glaucoma risk. This was conducted to determine whether these associations reflected potential causal effects rather than confounding or reverse causation. The MR analysis revealed causal relationships between foveal morphology and AMD risk, but not glaucoma risk. Specifically, the findings for pit volume were suggestive of a causal contribution to AMD risk (inverse-variance weighted [IVW] p = 0.03; OR per SD 38.81; 95% CI 1.32 –1142.30) (Supplementary Figs.16 and 17 and Supplementary Table 8). Although the confidence interval around this estimate was wide (limiting the precision with which the effect size could be estimated), it excluded the null and therefore remained consistent with a causal relationship under the assumptions of the Mendelian randomisation framework. This result was supported by a significant MR Egger (p = 0.049). Leave-one-out analysis indicated that four variants (Supplementary Table 9) had notable influence on the findings; however, none of these variants were identified as an outlier by Radial MR or found to be associated with confounding variables in the Open Targets resource.^26^

Recognising that our analyses may be impacted by the correlated nature of foveal traits, we conducted multivariable MR considering the causal relationship between all six foveal traits and AMD. Analysis of the multivariable model revealed significant risk of pleiotropy (Q-statistic p-value = 4.0 × 10^−8^) and evidence of weak instruments for pit volume (F-statistic = 3.92), mean slope (F-statistic = 2.08), rim radius (F-statistic = 2.59) and pit depth (F-statistic = 4.56). Overall, the multivariable analysis (using conventional IVW and Q-statistic minimization) was likely underpowered, limiting the ability to detect significant associations.

In summary, these findings provide evidence consistent with a causal contribution of pit morphology to AMD risk. Although the univariate MR results suggest that the leading trait is pit volume, the correlation between foveal parameters and the weak instrument bias in multivariable MR means that it remains possible that other foveal morphological traits could be important. We did not identify evidence supporting a causal relationship between foveal morphology and primary open-angle glaucoma; this suggests that the observational associations may reflect shared risk factors, confounding, or downstream consequences of disease, rather than a direct effect of foveal morphology on glaucoma risk. Notably, in all analyses, there was no evidence of reverse causation. Given that the results were not significant across all robust tests, they should be interpreted in the context of the assumptions described in the Methods section (and may be sensitive to violations of these assumptions).

## DISCUSSION

In the present study, we performed large-scale phenotypic and genetic analyses to better understand the variability and determinants of foveal morphology. By leveraging retinal imaging data from UK Biobank participants and a comprehensive foveal trait calculation tool (RETIMAT), we were able to detect differences in foveal morphology influenced by sex, genetic ancestry and genetic factors. We also found a notable association between foveal pit volume and AMD. Given the strong correlations among foveal traits, it is difficult to determine the extent to which the association with AMD is specific to pit volume.

We first analysed how foveal architecture varies at the population level. As the number of subjects in this study is an order of magnitude greater than in most prior reports, we were able to identify previously unrecognized variability in foveal pit morphology (e.g. 15-fold for pit volume). Sex and ancestry were found to significantly influence foveal morphology, an observation that is in keeping with the findings of previous smaller-scale studies.^7,9,14,15^ In line with previous research, males exhibited greater rim height, shorter rim radii and a steeper mean slope.^15^ Similarly, we found that individuals with African ancestries had wider, deeper and more voluminous foveal pits than their European counterparts, in line with findings by Zouache *et al.*^7^ The smaller East- and South-Asian ancestral groups fell between these two, consistent with the limited normative datasets available for Asian populations.^27,28^ It is highlighted that, in contrast to most previous studies, we used genetic ancestry, a metric that is more accurate and objective than ethnicity or self-reported ancestry.^29^ We also performed additional analyses to gain insights into the extent to which the observed difference between continental groups is driven by pigmentation. Our observations suggest that genetic ancestry tends to be an independent contributor irrespective of fundus pigmentation, especially for foveal pit volume and rim radius.

We found that a larger pit volume is a key risk factor for AMD, and that there is a 33% increase in risk per 0.1 mm^3^ increase in pit volume. This observation was supported by two independent approaches: Cox regression and Mendelian randomisation. The results of the latter substantially reduce the risk that the observed association is due to reverse causation or confounding, and support the assertion that pit volume is a risk factor for AMD rather than a correlate. The hazard ratio per one SD was 1.09 (95% CI 1.04–1.16) and the corresponding odds ratio (OR) per SD was 1.10 (95% CI 1.06–1.17) (Fig.5, Supplementary Tables 5 and 10). We note that several environmental and genetic risk factors have been previously associated with AMD, including: age (OR 1.1 per year), gender (OR 1.6), smoking (OR 1.9)^30^, *CFH* gene variants (e.g. homozygosity for rs1061170 corresponds to OR of 4.7)^31^, *ARMS2* locus variants (e.g. homozygosity for rs10490924 corresponds to OR of 8.6)^32^ and family history (OR 10.8).^33^ Although AMD selectively affects the foveal/macular region, to date, anatomical risk factors have been understudied^9^ and, to our knowledge, this is the first time that specific foveal morphological parameters are found to set the stage for susceptibility to AMD. It can be speculated that the link between a larger pit and AMD risk is driven by differences in the distribution of macular xanthophyll pigments and/or by alterations in photoreceptor cell densities in the vulnerable parafoveal area of low rod-to-cone ratio.^34^ Future advanced ophthalmic imaging and mathematical modelling studies will provide further insights into how foveal anatomy predisposes to this leading cause of visual impairment.

Following phenotypic analyses, including of foveal traits that have received little attention so far (such as foveal volume and radius), we performed genetic association studies. We found that the observed morphological diversity has a strong genetic underpinning, with a high level of heritability across most traits (Supplementary Table 6). Pigmentation-related genes such as *TYR* and *TSPAN10* surfaced across multiple foveal traits, underscoring the role of melanin synthesis in the shared developmental pathways that shape foveal morphology (Table 2). This is in keeping with previous observations in albinism^35,36^ and in a smaller cohort where a link between the common functional *TYR:*c.1205G>A (p.Arg402Gln) [rs1126809] variant and decreased rim radius was reported.^37^

While many of the lead variants detected here have previously been linked to retinal traits, at least 130 of them have no such prior associations (Supplementary Data). Examples include *CYP1A1* and *KMT2A. CYP1A1* (lead marker: rs200391170; association with mean slope; p = 2.2 × 10^−21^) encodes a major cytochrome P450 enzyme and is involved in the metabolism of both endogenous substances and environmental chemicals. It has been implicated in the conversion of all-trans retinol to all-trans retinoic acid in human foetal tissues.^38^ Also consistent with a role for retinoic acid signalling, we identified a variant associated with pit volume upstream of *CYP26C1* (lead marker: rs35357127; p = 5.1 × 10⁻^15^; Supplementary Data). *CYP26C1* encodes a retinoic acid-degrading enzyme and its chick orthologue has been linked to foveal patterning through local modulation of retinoic acid levels.^39,40^ *KMT2A* (lead marker: rs7948661; association with rim radius; p = 5.9 × 10^−10^) encodes a histone methyltransferase that mediates H3K4 methylation, a key epigenetic modification associated with transcriptional activation.^41^ The relevant molecule regulates the expression of developmental genes across various tissues, including the developing retina, and conditional *Kmt2a* knockout in mice has been shown to lead to disrupted retinal layering.^42,43^

A subset of the detected lead variants (n=17) has been previously associated with retinal thickness measurements (either clinically-used^16–18^ or deep learning based^19–21^) but not with foveal/macular traits (Supplementary Data). These novel associations reported here enhance the interpretability of the relevant signals and allow more robust links to be drawn with central retinal development/patterning. Notable examples include *FGFR2* and *SPRY2. FGFR2* (lead marker: rs146727842; association with mean slope; p = 1.4 × 10^−20^) encodes a receptor tyrosine kinase that is essential for embryonic development.^45^ *FGFR2* has been shown to interact with *FGF8*, whose expression is required to pattern the chick fovea-like region.^39,40,46^ *SPRY2* (lead marker: rs7996082; association with rim radius; p = 9 × 10^9^) encodes a member of the Sprouty family of proteins, which act as negative regulators of the FGF signalling pathway.^47^ Experimental evidence from murine models suggests a prominent role for *SPRY2* in eye development.^48–50^

This study has several limitations. First, obtaining accurate quantitative measurements of OCT-derived retinal features is inherently challenging. One issue is image scaling, as precise measurements require knowledge of the exact lateral scale of each scan. To address lateral magnification, we included spherical equivalent refractive error as a covariate in our analyses. This is a pragmatic alternative to using ocular axial length^33,51^ (which is not available within UK Biobank) but it is less precise and does not fully account for individual variation in retinal image scaling. Our analytical framework nonetheless closely aligns with most previous genetic association studies involving retinal imaging-derived phenotypes^16,19–21^ and, reassuringly, the findings of our phenotypic analyses showed good agreement with those from independent, smaller, more targeted studies.^52,53^ To evaluate whether the use of refractive error in this context influenced our time-to-event results, we performed sensitivity analyses excluding participants at increasingly strict spherical equivalent thresholds (±6, ±2, ±1 dioptres). The hazard ratios remained stable throughout (Supplementary Table 11). Second, our image quality control process included removal of extreme values for foveal morphological measures; while this approach helps to exclude failed OCT image segmentations, it may also result in the exclusion of true anatomical extremes. Third, although our analyses were adjusted for established covariates known to influence foveal morphology, additional unaccounted-for covariates and environmental factors may still contribute to residual confounding. Fourth, our primary analysis used a single, prespecified eye per participant. Alternative approaches, including bilateral averaging, are also valid and future studies could examine the consistency of genetic findings across phenotype definitions. Fifth, the internal validation analysis assessed overall concordance between discovery and validation effect estimates but did not assess validation at the level of independent signals (one variant per locus). Sixth, we relied on UK Biobank healthcare record data to identify cases of AMD and glaucoma, and to exclude relevant cases at baseline. These records are known to be incomplete and several alternative approaches using retinal imaging data have been proposed.^54,55^ Although such methods have shown positive diagnostic performance, they can overestimate disease prevalence,^55^ and we opted to use a conventional record-based case definition. We utilised the H35 ICD-10 code as a proxy of ‘AMD cases’, consistent with its use in some previous studies.^56,57^. However, as described in the Methods, this code encompasses a broad group of retinal disorders in addition to AMD, and some degree of diagnostic heterogeneity remains possible. Reassuringly, the Mendelian randomisation analyses provide support for the interpretation that the reported relationships are driven by AMD-related pathology. Seventh, while the current study leverages population-scale genetic and epidemiological approaches to investigate associations between foveal morphology and ocular disease, it is not designed to establish the underlying biological mechanisms, which will require targeted mechanistic follow-up. Finally, while the UK Biobank was designed to be broadly representative of the ageing UK population, it is subject to selection biases. In addition, the limited representation of non-European ancestry groups further constrains generalisability. All reported associations should be interpreted in light of these limitations.^58,59^

In conclusion, we show that (i) foveal architecture is patterned by sex, genetic ancestry and genes related to pigmentation and (ii) variation in foveal morphology, including pit volume, is associated with AMD risk. These findings suggest that foveal morphology is biologically relevant to macular vulnerability and invite mechanistic work to dissect how developmental pathways intersect with ageing to drive retinal degeneration.

## METHODS

### Ethical approval

The UK Biobank has received approval from the UK National Information Governance Board for Health and Social Care and the National Health Service North West Centre for Research Ethics Committee (Ref: 11/NW/0382). This research was conducted using the UK Biobank Resource under projects 53144 and 49978. All participants provided informed consent to participate in the UK Biobank study. All investigations were conducted in accordance with the tenets of the Declaration of Helsinki.

### UK Biobank: cohort characteristics and ophthalmic phenotyping

We used data from the UK Biobank, a resource containing genomic and health information from 502,355 individuals from across the United Kingdom, aged between 40 and 69 years at the time of recruitment.^3^ Subsets of UK Biobank volunteers underwent enhanced phenotyping including visual acuity testing (>130,000 individuals), non-cycloplegic autorefraction (>125,000 individuals) and imaging of the central retina using colour fundus photography and OCT (>84,000 individuals). Baseline ophthalmic assessment was conducted between 2009 and 2010 for most participants and between 2012 and 2013 for a smaller subset.^4^

Best-corrected visual acuity was assessed using a computer-displayed LogMAR chart (Precision Vision, LaSalle, IL, US) (UK Biobank data-fields: 5208 and 5201). Participants wore their habitual distance correction and were tested at 4m; if unable to identify any letters, the chart was moved to 1m. Starting from the top line, testing stopped once ≥2 letters on a line were misread.^4^

Non-cycloplegic autorefraction was carried out using a Tomey RC 5000 device (Tomey Corp., Nagoya, Japan). The spherical equivalent refractive error (spherical error + 0.5 × cylindrical error) was subsequently calculated for each participant (data-fields 5084-5085; 5086-5087).^4^

Colour fundus photographs and spectral-domain OCT images were acquired using the Topcon 3D-OCT 1000 Mark II device (Topcon Corp., Tokyo, Japan), without pharmacological dilation (data-fields 21015, 21016, 21011, 21013). OCTs were obtained using a ‘3D macular volume’ protocol (6 × 6 mm; 128 horizontal B-scans, each comprising 512 A-scans arranged in a raster pattern).^5,44,60^ The right eye was imaged first. Our analyses predominantly focused on left eye images as familiarity with the test generally yielded higher-quality scans.^5,19^ A prespecified eye was used to avoid the reduced sample size associated with bilateral averaging and the additional modelling assumptions required for joint analysis of both eyes. Prespecified-eye analyses have been adopted in several previous ophthalmic imaging and genetic studies,^17,19,61–66^ although alternative approaches, including bilateral averaging, have also been widely used.^16,18,20,21,67^ For robustness, an average value for each foveal trait (calculated using both left and right eye measurements but falling back to the single available eye when only one scan passed QC) was also used in key phenotypic analyses (Supplementary Fig.1 and 3). Comparisons of foveal traits in the left, right and averaged datasets are presented in Supplementary Table 12).

### Fundus pigmentation quantification

We used a recently described method^22^ to evaluate fundus pigmentation in UK Biobank participants. This involved calculating the Retinal Pigment Score (RPS), a continuous quantitative metric that is thought to primarily reflect choroidal pigmentation.^59^

The RPS was derived using the approach described by Rajesh *et al*. which entails selecting fundus photographs of sufficient quality (*i.e.* those labelled as ‘Good’ by Automorph, an established deep learning tool)^68^ and processing them through the RPS pipeline to quantify background pigmentation.^22^ It is noted that the RPS is designed to quantify background pigmentation while reducing the influence of image illumination, by leveraging colour features defined in the CIELAB colour space. ^22^

### OCT quality control and foveal morphological parameter extraction

OCT scans were filtered for overall quality in line with previous studies.^19,21^ Briefly, we discarded OCTs that (j) had a quality score below 40 and (ii) fell within the lowest 10% of the inner limiting membrane (ILM) edge-strength indicator (a metric that captures the sharpness and continuity of the ILM boundary and flags blinks, signal dropouts and major segmentation errors).^21^ Where multiple scans were available per instance, only the primary (index 0) image was used as quality metrics are not provided for additional scans.

OCT images meeting the above quality control criteria were processed using RETIMAT, an open-source MATLAB toolbox for OCT image analysis. The processing pipeline included the following steps:

- parsing Topcon (.fda) files to extract the ILM and Bruch’s membrane (BM) surfaces delineated by the device software;
- computing total retinal thickness maps;
- automatically aligning thickness maps to the foveal centre;
- transforming raster thickness maps into a radial pattern; and
- computing morphometric values.

RETIMAT has been used successfully in previous studies and its fully automated methods for fovea location and analysis have been independently assessed, making it well-suited for high-throughput analyses.^15,69^ Notably, RETIMAT functions primarily as a utility rather than as a segmentation algorithm. It does not perform any image segmentation or grading itself, and instead operates entirely on the ILM–BM boundaries supplied by the Topcon device. These particular boundaries are among the most robust OCT features to segment due to their distinct intensity gradients. We are not aware of evidence that factors such as fundus pigmentation materially influence OCT signal quality or segmentation performance (among scans that meet standard quality-control criteria). Nonetheless, as with any imaging-derived measurement, a degree of residual technical variation cannot be excluded, and this remains an area for further investigation.

We focused on six RETIMAT-derived foveal traits: foveal pit volume, rim radius, rim height, mean slope, pit depth and central foveal thickness (CFT) (Fig.1, Table 1). Aiming to study the phenotypic variability of these parameters, we assembled a broad ‘phenotypic analysis’ cohort. This included unrelated UK Biobank participants: whose OCT scans passed the imaging quality control criteria for this study; whose RETIMAT-derived foveal trait values did not fall in the 1% upper or lower extreme; and who were assigned to one of the four main genetically-inferred ancestral groups (EUR, AFR, SEA and SAS – see ‘Genetic ancestry assignment’ section below). Aiming to study the genetic architecture of foveal traits, a second, more focused ‘genetic analysis’ cohort was defined. This was limited to unrelated participants of European-like ancestries with: OCT scans; no sex chromosome aneuploidy; and not flagged as outliers for heterozygosity nor missingness. Individuals with RETIMAT-derived foveal trait values exceeding 2.5 SD were also excluded (Supplementary Fig.18); this cut-off was selected in accordance with the approach used in previous genetic association studies of retinal thickness measurements.^16,19–21^ The final GWAS sample comprised 36,205 unrelated individuals with European-like ancestries (29,710 primary plus 6,495 internal validation); the broader phenotypic cohort totalled 39,521 individuals across ancestries.

### Genetic ancestry assignment

To facilitate inter-ancestry analyses of foveal traits, we sought to characterize the genetic ancestry of UK Biobank participants. A subset of 409,373 participants with European-like ancestries is readily available (data-field 22006) and we aimed to obtain genetic ancestry estimates for the remaining 78,296 individuals. Using a previously described approach^70^ we projected these UK Biobank participants onto reference genetic data from the 1000 Genomes Project.^71^ In brief, directly genotyped single-nucleotide variants from UK Biobank were matched to those in the 1000 Genomes Project (Phase 3, v5a.20130502) and merged using PLINK v2.0,^72^ retaining 718,487 high-quality variants. Linkage disequilibrium (LD) pruning (--indep-pairwise 50 10 0.025; 50-variant window advanced in 10-variant steps, r² < 0.025) was performed using LD estimates from unrelated 1000 Genomes YRI individuals. We additionally excluded 24 regions of long-range LD, yielding 30,320 independent variants. Ancestry proportions were estimated using ADMIXTURE (v1.3.0),^73^ with four 1000 Genomes reference populations: GBR (European, EUR), YRI (African, AFR), ITU (South Asian, SAS) and CHS (East Asian, EAS). To avoid potential confounding from heterogeneous admixture in the Americas, no Admixed American (AMR) reference population was included; consequently, no participants were assigned AMR ancestry. Individuals were retained for downstream phenotypic analyses if ≥80% of their inferred ancestry was assigned to one of the four reference groups.

### Statistical analyses of foveal phenotypes

We characterized the independent contributions of genetic ancestry, sex, refractive error and age (at OCT acquisition) to variability in foveal morphological traits by fitting separate multivariable linear regression models for each foveal trait. Each trait was standardized (Z-scored) prior to modelling so that the regression coefficients represented the change in the trait in standard deviation (SD) units per one unit of the predictor. Regression coefficients, 95% confidence intervals and p-values were subsequently extracted.

To quantify the contributions of fundus pigmentation (RPS) and genetic ancestry to variability in foveal morphology, we fitted linear regression models for each trait, as follows: The two predictors were analysed together because pigmentation traits vary across ancestry groups and may reflect overlapping sources of variation.

*Model 1:* included RPS, age at OCT acquisition, spherical equivalent refractive error and sex.

*Model 2* included genetic ancestry (as a categorical variable), age at OCT acquisition, spherical equivalent refractive error and sex.

*Model 3*: included both RPS and genetic ancestry terms, along with age at OCT acquisition, spherical equivalent refractive error and sex.

From each model, we extracted adjusted R² values as a measure of model fit. Changes in adjusted R² were calculated by comparing the full model against models excluding either RPS or ancestry terms, allowing comparison of the relative impact of these predictors on model fit across the six foveal traits.

Finally, the association between foveal morphology and visual acuity was studied using a linear regression approach with age, sex, spherical equivalent refractive error and genetic ancestry used as covariates.

### Genome-wide association studies (GWAS)

We performed a common-variant GWAS using an additive linear model in REGENIE v3.1.1.^23^ To meet the normality assumptions of GWAS, we applied rank-based inverse normal transformation to any foveal traits that deviated from a normal distribution. For this analysis, we focused only on individuals of European-like genetic ancestries (data-field 22006). We performed a primary GWAS and validated the findings in an independent, temporally distinct subset of the same cohort. The focus of the former was on the 67,250 individuals that were imaged at the time of their baseline visit (Instance 0, ‘Initial assessment visit (2006–2010)’). The focus of the internal validation study was on the 15,568 different participants that were imaged for the first time during their first repeat assessment (Instance 1, ‘First repeat assessment visit (2012–2013)’. Thus, the two analyses were performed on non-overlapping samples.

To process the imputed genotype data, we applied conventional quality control filters using PLINK2.^72^ Variants were retained if they had: an imputation quality score above 0.8; a minor allele frequency (MAF) of 5% or more; and a minor allele count (MAC) of 20 or more. Variants with a Hardy-Weinberg equilibrium p-value below 1 × 10^−15^ were excluded, along with those with missingness greater than 10%. Individuals with more than 10% missing genotypes^23^ or with genetic kinship to other UK Biobank participants (based on data-field 22021) were also excluded.

The GWAS model was adjusted for age at recruitment (data-field 21022), sex (data-field 31), height (data-field 50), weight (data-field 21002), spherical equivalent refractive error (calculated from data-fields 5085 and 5086 and defined as spherical error + 0.5 × cylindrical error) and the first 20 genetic principal components (data-field 22009). Genome-wide statistical significance was set at p < 8.3 × 10^−9^ after correcting for multiple testing across the six studied foveal traits.

To refine the obtained association signals, further analyses were performed using the GCTA-COJO tool (conditional and joint multiple-variant analysis).^24^ Variants located on different chromosomes, or more than 10 Mb apart, were treated as unlinked. Lead genetic variants were then annotated using Ensembl,^74^ Open Targets,^26,75^ and GWAS Catalog^76^ data (accessed on June 24, 2025). To aid interpretation of the strongest signals, LD relationships between lead variants identified by GCTA-COJO that were located within 1 Mb of one another were inspected using the LDlink tool.^77^

### Rare variant burden testing

Rare variant analyses were performed using REGENIE (v2.2.4)^23^ on the UK Biobank OQFE exome dataset (data-field 23157), comprising ∼12.3 million coding variants. Following quality control and annotation filtering, ∼2.7 million rare variants were retained.

Gene-based burden testing was conducted across 18,108 protein-coding genes evaluated in the UK Biobank exome dataset. Variant annotation and deleteriousness scoring were obtained from the UK Biobank OQFE helper files. Each variant was assigned to a gene and a functional class, including loss-of-function (LoF), missense variants (sub-classified by the number of five in silico algorithms predicting deleteriousness: missense(0/5), missense(≥1/5) and missense(5/5)) and synonymous variants.

Rather than pre-filtering variants based on predicted deleteriousness, we constructed inclusive gene-level rare variant burdens by aggregating all rare (AAF ≤ 0.01) protein-altering variants within each gene. This included LoF and all missense categories, thereby capturing the full spectrum of rare coding variation without relying on *in silico* prediction thresholds.

The same six foveal traits and covariates used in the common-variant GWAS were applied in the rare variant analyses, with rank-based inverse normal transformation applied to non-normally distributed traits. Gene-level association results were summarised and assessed for statistical significance using Bonferroni correction for multiple testing across 18,108 genes and six traits (p < 4.60 × 10^−7^).

### LD score regression: heritability estimation and global genetic correlation analysis

The global genetic correlations between the six studied foveal traits (pit volume, rim radius, rim height, mean slope, pit depth and CFT) and RPS were calculated using LD score regression.^78^ The GWAS summary statistics for RPS were obtained from a study by Julian *et al.* (GWAS Catalog GCST90672040).^61^ The analysis adhered closely to the methodology described by Bulik-Sullivan *et al.*,^78^ with genetic variants filtered according to the following criteria: inclusion in HapMap3; minor allele frequency > 0.01; imputation score > 0.9; exclusion of strand-ambiguous variants; and removal of duplicated variants. LD scores were derived from the European subset of the 1000 Genomes Project dataset.^71^

LD score regression was also used to calculate heritability, λG (genomic inflation factor), intercept and ratio (denoting the proportion of inflation resulting from confounding or bias) for each of the six studied foveal traits.

### Cox regression analysis

We performed Cox proportional hazards regression to investigate the association between foveal traits and incident age-related macular degeneration (AMD) and glaucoma. Participants with available OCT-derived foveal traits and outcome data were included in the analysis (n = 39,521). Disease status was defined using hospital inpatient ICD-10 diagnosis fields (data-field: 41202) together with the corresponding First Occurrence algorithm fields (data-field 131182 for H35-related outcomes and data-field 131186 H40-related outcomes) to ensure complete capture of the earliest recorded diagnosis date.

The ICD-10 code H35 encompasses a heterogeneous group of retinal disorders, including “retinopathy of prematurity”, “other proliferative retinopathy”, “degeneration of macula and posterior pole”, “peripheral retinal degeneration”, “separation of retinal layers” and “hereditary retinal dystrophy”, among others. In our analysis, disease events were defined prospectively (post-imaging), which excludes several conditions within H35 that are congenital or typically diagnosed earlier in life (e.g. retinopathy of prematurity, hereditary retinal dystrophies). In addition, OCT imaging captures central (macular) rather than peripheral retinal structure, making it unlikely that conditions primarily affecting the peripheral retina would influence the foveal metrics examined here. Taken together, these considerations suggest that associations between foveal metrics and the H35 ICD-10 code are likely to be driven predominantly by AMD, a central retinal disorder that commonly emerges in the age group studied. We further evaluated this assumption using an AMD-specific Mendelian randomisation analysis, as described in the next section.

Disease onset was defined as the earliest recorded date of an AMD- or glaucoma-related ICD-10 code. Participants were censored at the last available follow-up date (May 31, 2022), if their imaging date was missing, or if disease onset occurred before the scan date. By the censor date, the mean age of participants was 69 years, placing the cohort well within the typical age range for AMD. As our focus was incident disease, individuals with a recorded AMD diagnosis at or before baseline were excluded. AMD severity was not included as a covariate because it is only defined among those who already have the disease and therefore cannot inform models of AMD onset.

Cox regression models were fitted using the ‘lifelines’ Python package,^79^ with time-to-event (in days) as the outcome and foveal traits as the predictors. All models were adjusted for age at image acquisition, sex, spherical equivalent refractive error and genetic ancestry. Ancestry was modelled using one-hot encoded genetic ancestry categories with individuals of European-like ancestries serving as the reference group. Models were fit for each trait–disease pair. Hazard ratios (HR), 95% confidence intervals and p-values were extracted. The p-values were corrected for multiple testing using the Benjamini–Yekutieli (BY) false discovery rate (FDR) method. The proportional-hazards assumption was not formally assessed for these models.

To account for potential imprecision introduced by using spherical equivalent rather than axial length, we conducted sensitivity analyses to assess the influence of extreme refractive error. Individuals with spherical equivalent values beyond ±6, ±2 or ±1 dioptres were excluded in separate Cox regression models.

As a complementary cross-sectional analysis, we performed logistic regression to test whether foveal traits measured at the time of imaging were associated with disease status. In contrast to the incident-event Cox analyses, cases were defined as individuals with a recorded diagnosis of the relevant condition before the censoring date (May 31, 2022, or date of death if earlier), irrespective of whether diagnosis occurred before or after imaging. Participants who did not meet the case definition were treated as controls. Covariates included age at image acquisition, sex, spherical equivalent refractive error and genetic ancestry (encoded as dummy variables). Analyses were run separately for each foveal trait and the logistic regression models were fitted using the ‘statsmodels’ Logit function in Python^80^. Odds ratios (with 95% confidence intervals) and p-values were extracted.

### Causal inference using two-sample Mendelian randomisation

Potential causal associations between AMD, glaucoma and the studied foveal traits were assessed using univariable two-sample Mendelian randomisation (MR).^81,82^ AMD was instrumented using a mixed (combined dry and wet) AMD GWAS by Han *et al*. (full summary statistics were obtained directly from the study authors; publicly available top associations: GWAS Catalog GCST010284).^83^ Glaucoma was instrumented using a primary open-angle glaucoma GWAS (excluding UK Biobank participants) by Gharahkhani *et al.* (GWAS Catalog GCST90011767).^84^ Population overlap biases univariable two-sample MR in the direction of causation. Accordingly, the populations contributing toward traits determined to be causal were inspected to ensure that there was no overlap. Genetic instruments were selected based on p < 5 × 10^−8^ in relation to each exposure trait. Where an exposure instrument was not present in the outcome dataset, we sought to identify a suitable proxy. Proxy variants were single-nucleotide variants that were in high LD with exposure-associated variants which were not present in the outcome dataset. Proxies were identified using the Ensembl server,^74^ selecting variants with an LD R^2^ value of ≥0.9. Genetic variants were clumped using an LD R^2^ value of 0.001 and a genetic distance cut-off of 10,000 Kb. This was achieved using the 1000 Genomes Project European superpopulation reference panel and the TwoSampleMR PLINK clumping function.^71,72^ The effects of instruments on outcomes and exposures were harmonized to ensure that the β values (*i.e.* the regression analysis estimates of effect size) were signed with respect to the same alleles. For palindromic alleles (*i.e.* alleles that are the same on the forward as on the reverse strand), those with minor allele frequency (MAF) > 0.42 were omitted from the analysis to reduce the risk of errors due to strand issues. In addition to using a range of Mendelian randomisation methods and quality control measures (as detailed below), we endeavoured to remove pleiotropic instruments and outliers from the analysis. To achieve this, instruments that were more statistically significant (in terms of p-value) for the outcome than the exposure were removed. Radial Mendelian randomisation,^85^ an approach that detects outlying instruments, was also used for outlier identification, and outliers were subsequently removed.^85^ The Cochran’s Q test was performed for each analysis. The MR Egger intercept^86^ was used to detect horizontal pleiotropy. The I^2^ statistic was calculated as a measure of heterogeneity between variant specific causal estimates. An I^2^ < 0.9 indicates that MR Egger is more likely to be biased towards the null through violation of the “NO Measurement Error” (NOME) assumption. Our primary analysis was a multiplicative random effects inverse variance weighted (IVW) test, which is the most efficient analysis method with valid instrumental variables.^87^

Mendelian randomisation relies on several assumptions for valid causal inference. These are (i) the instrumental variables (single-nucleotide polymorphisms) are associated with the exposure; (ii) the instrumental variables are not associated with confounders of the exposure-outcome relationship; and (iii) the instrumental variables influence the outcome only through the exposure of interest (*i.e.* no horizontal pleiotropy).^87^

Robust Mendelian randomisation methods (‘tests’), which can provide valid causal inference under weaker assumptions than IVW analysis, were also applied. These included weighted median (robust to up to 50% of invalid instruments), weighted mode (robust to >50% of invalid instruments but assumes more variants estimate the true causal effect than estimate any other quantity), MR Egger (robust to pleiotropic effects of variants, provided the effects are independent of the variant-exposure associations) and a leave-one-out analysis using the MRE IVW method (to test for highly influential variants).^86–89^

When appraising the results of a Mendelian randomisation study, the confidence with which one can assert causality is influenced by both the strength of association in the primary analysis (IVW) and the range of robust tests in which the results are consistent. Where the IVW estimate is significant but robust tests are not, the findings may still be in keeping with a causal relationship; however, they rely more heavily on the core assumptions of Mendelian randomisation and therefore warrant more cautious interpretation.

Consistent with methodological recommendations for Mendelian randomisation,^87^ we systematically evaluated all empirically testable assumptions underpinning our analyses. This included inspection of instrument strength, assessment of heterogeneity using Cochran’s Q, evaluation of directional horizontal pleiotropy via the MR-Egger intercept and examination of the NOME assumption using the I² statistic. We additionally performed radial MR to identify disproportionately influential variants, removed instruments with stronger associations with the outcome than the exposure and carried out leave-one-out analyses to confirm that results were not driven by single variants. Together, these diagnostics provide a comprehensive assessment of instrument validity and the plausibility of the underlying Mendelian randomisation assumptions.

### Statistics and reproducibility

No statistical methods were used to predetermine sample size; all participants passing quality control were included. Phenotypic analyses used 39,521 UK Biobank participants. Genetic analyses used 36,205 unrelated participants of European-like ancestry, split into a primary discovery sample (n=29,710) and a non-overlapping internal validation sample (n=6,495). Linear regression was used to assess associations between foveal traits and demographic/ophthalmic covariates. Cox proportional hazards models were used for time-to-event analyses, with Benjamini–Yekutieli FDR correction for multiple testing. GWAS was performed using REGENIE (study-wide threshold p < 8.3 × 10⁻⁹). Heritability and genetic correlations were estimated using LD score regression. Mendelian randomisation used IVW regression as the primary method, with weighted median, weighted mode and MR-Egger as sensitivity analyses. Further detail on each method is provided in the sections above.

## Data availability

The UKB dataset is available under restricted access through the procedure described by UK Biobank at https://www.ukbiobank.ac.uk/use-our-data/. GWAS summary statistics for AMD and primary open-angle glaucoma were obtained from Han *et al.*^83^ and Gharahkhani *et al.*^84^ respectively. GWAS summary statistics generated in this study will be deposited in the GWAS Catalog^76^. Source data for Fig. 2 are provided in the Supplementary Data file and in Supplementary Tables 2 and 3. Source data for Fig. 2 are provided in the Supplementary Data file and in Supplementary Tables 2 and 3. Source data for Figs. 3, 4, 5 and 7 are provided in Supplementary Tables 1, 4, 5 and 7, respectively.

## Code availability

The scripts used to analyse the datasets included in this study are available at https://github.com/davidjohngreen/fovea/ archived at ^90^ under an MIT License. Foveal morphological parameters were extracted from UK Biobank OCT images using RETIMAT, an open-source toolbox available at https://github.com/mu-biomed/retimat. Analyses requiring UK Biobank participant-level data as input cannot be run without independently obtaining approved UK Biobank data access, as this data is not included in the repository and cannot be redistributed by the authors (https://www.ukbiobank.ac.uk/use-our-data/).

## ACKNOWLEDGEMENTS

We thank the participants and coordinators of the UK Biobank study for their contributions and commitment, which made this research possible. This research was conducted using the UK Biobank Resource under projects 53144 and 49978

## FUNDING STATEMENT

We acknowledge the following sources of funding: the Wellcome Trust (224643/Z/21/Z, Clinical Research Career Development Fellowship to P.I.S.); the UK National Institute for Health Research (NIHR) Clinical Lecturer Programme (CL-2017-06-001 to P.I.S.); the NIHR Manchester Biomedical Research Centre (NIHR 203308 to P.I.S. and G.C.B.); the EMBL European Bioinformatics Institute (EMBL-EBI) (S.T., T.F. and E.B.); Medical Research Council (MRC) Clinical Research Training Fellowship (MR/Z504105/1, to T.H.J), and the Howard Hughes Medical Institute (C.L.C). Research reported in this publication was supported in part by the National Eye Institute of the National Institutes of Health (NIH) under award numbers R01EY017607 and R01EY033580. The content is solely the responsibility of the authors and does not necessarily represent the official views of the NIH.

The UK Biobank Eye and Vision Consortium is supported by funding from the NIHR Biomedical Research Centre at Moorfields Eye Hospital and UCL Institute of Ophthalmology, the Alcon Foundation and the Desmond Foundation

## AUTHOR CONTRIBUTIONS STATEMENT

P.I.S., D.J.G. and T.H.J. conceived and designed the experiments. The UK Biobank Eye and Vision Consortium, D.R., H.N.V.J., U.A., M.B., J.E.S., G.C.B., T.F., E.B., C.L.C., J.C. and P.I.S. provided datasets and analytical tools. D.J.G., D.R., T.H.J., S.T., H.N.V.J., T.F., C.L.C. and P.I.S. analysed the data. P.I.S. and D.J.G. wrote the manuscript with support from T.H.J.. All authors critically revised and approved the manuscript.

## COMPETING INTERESTS STATEMENT

E.B. is a paid consultant and equity holder of Oxford Nanopore, a paid consultant to Dovetail and a non-executive director of Genomics England, a limited company wholly owned by the UK Department of Health and Social Care. All other authors declare no competing interests. D.R. is an employee of RetinAI, but performed this work independently of RetinAI.

## UK Biobank Eye and Vision Consortium

Naomi Allen¹³, Tariq Aslam¹⁴, Denize Atan¹⁵, Konstantinos Balaskas¹⁶, Sarah Barman¹⁷, Jenny Barrett¹⁸, Paul Bishop¹⁴, Graeme Black¹⁴, Tasanee Braithwaite¹⁹, Roxana Carare²⁰, Usha Chakravarthy²¹, Michelle Chan¹⁶, Sharon Chua²², Alexander Day¹⁶, Parul Desai¹⁶, Bal Dhillon²³, Andrew Dick¹⁵, Alexander Doney²⁴, Cathy Egan¹⁶, Sarah Ennis²⁰, Paul Foster²², Marcus Fruttiger²², John Gallacher¹³, David Garway-Heath²², Jane Gibson²⁰, Jeremy Guggenheim²⁵, Chris Hammond²⁶, Alison Hardcastle²², Simon Harding²⁷, Ruth Hogg²¹, Pirro Hysi²⁶, Peng Tee Khaw²², Anthony Khawaja¹⁶, Gerassimos Lascaratos¹⁶, Adam Lewandowski¹³, Thomas Littlejohns¹³, Andrew Lotery²⁰, Robert Luben²², Phil Luthert²², Tom Macgillivray²³, Sarah Mackie¹⁸, Savita Madhusudhan²⁸, Bernadette Mcguinness²¹, Gareth Mckay²¹, Martin Mckibbin²⁹, Tony Moore²², James Morgan²⁵, Eoin O’Sullivan³⁰, Richard Oram³¹, Chris Owen³², Praveen Patel¹⁶, Euan Paterson²¹, Tunde Peto²¹, Axel Petzold³³, Nikolas Pontikos²², Jugnoo Rahi³⁴, Alicja Rudnicka³², Naveed Sattar³⁵, Jay Self²⁰, Sobha Sivaprasad¹⁶, David Steel³⁶, Irene Stratton³⁷, Nicholas Strouthidis¹⁶, Cathie Sudlow²³, Zihan Sun²², Robyn Tapp³², Dhanes Thomas¹⁶, Emanuele Trucco²⁴, Adnan Tufail¹⁶, Ananth Viswanathan¹⁶, Veronique Vitart²³, Mike Weedon³¹, Katie Williams²⁶, Cathy Williams¹⁵, Jayne Woodside²¹, Max Yates³⁸, Yalin Zheng²⁷

¹³ University of Oxford, Oxford, United Kingdom

¹⁴ The University of Manchester, Manchester, United Kingdom

¹⁵ University of Bristol, Bristol, United Kingdom

¹⁶ Moorfields Eye Hospital, London, United Kingdom

¹⁷ Kingston University, London, United Kingdom

¹⁸ University of Leeds, Leeds, United Kingdom

¹⁹ St Thomas’ Hospital, London, United Kingdom

²⁰ University of Southampton, Southampton, United Kingdom

²¹ Queen’s University Belfast, Belfast, United Kingdom

²² UCL Institute of Ophthalmology, London, United Kingdom

²³ University of Edinburgh, Edinburgh, United Kingdom

²⁴ University of Dundee, Dundee, United Kingdom

²⁵ Cardiff University, Cardiff, United Kingdom

²⁶ King’s College London, London, United Kingdom

²⁷ University of Liverpool, Liverpool, United Kingdom

²⁸ Royal Liverpool University Hospital, Liverpool, United Kingdom

²⁹ Leeds Teaching Hospitals NHS Trust, Leeds, United Kingdom

³⁰ King’s College Hospital, London, United Kingdom

³¹ University of Exeter, Exeter, United Kingdom

³² St George’s, University of London, London, United Kingdom

³³ UCL Institute of Neurology, London, United Kingdom

³⁴ UCL Institute of Child Health, London, United Kingdom

³⁵ University of Glasgow, Glasgow, United Kingdom

³⁶ Newcastle University, Newcastle, United Kingdom

³⁷ Gloucestershire Hospitals NHS Foundation Trust, Gloucester, United Kingdom

³⁸ University of East Anglia, Norwich, United Kingdom

## Notes

### Competing Interest Statement

Ewan Birney is a paid consultant and equity holder of Oxford Nanopore, a paid consultant to Dovetail, and a non-executive director of Genomics England, a limited company wholly owned by the UK Department of Health and Social Care. All other authors declare no competing interests. David Romero-Bascones is an employee of RetinAI, but performed this work independently of RetinAI.

### Author Declarations

The UK Biobank has received approval from the UK National Information Governance Board for Health and Social Care and the National Health Service North West Centre for Research Ethics Committee (Ref: 11/NW/0382). This research was conducted using the UK Biobank Resource under projects 53144 and 49978.

### Summary of Updates

We refined the interpretation of the associations between foveal morphology and AMD, clarified limitations relating to the internal GWAS validation and Cox regression analyses, and updated data availability and source-data reporting.

