## Supplementary Material for "Genetic insights into foveal morphology and its associations with pigmentation and age-related macular degeneration"

### TITLE

### AUTHOR LIST & AFFILIATIONS

David J. Green[1], Thomas H. Julian[1,2-4], David Romero-Bascones[5], Sofia Torchia[4], Heer N.V. Joisher[6-8], UK Biobank Eye and Vision Consortium, Unai Ayala[5], Maitane Barrenechea[5], Jay E. Self[9,10], Graeme C. Black[1,11], Tomas Fitzgerald[4], Ewan Birney[4], Constance L. Cepko[6-8], Joseph Carroll[12], Panagiotis I. Sergouniotis[1,2,4,11]

[1] Division of Evolution, Infection and Genomics, School of Biological Sciences, Faculty of Biology, Medicine and Health, University of Manchester, Manchester, UK.

[2] Manchester Royal Eye Hospital, Manchester University NHS Foundation Trust, Manchester, UK.

[3] Christabel Pankhurst Institute, The University of Manchester, Manchester, UK

[4] European Molecular Biology Laboratory, European Bioinformatics Institute (EMBL-EBI), Wellcome Genome Campus, Cambridge, UK

[5] Biomedical Engineering Department, Faculty of Engineering (MU-ENG), Mondragon Unibertsitatea, Mondragón, Spain.

[6] Department of Genetics, Blavatnik Institute, Boston, MA, USA

[7] Department of Ophthalmology, Harvard Medical School, Boston, MA, USA

[8] Howard Hughes Medical Institute, Chevy Chase, MD, USA

[9] Clinical and Experimental Sciences, Faculty of Medicine, University of Southampton, Southampton, UK

[10] Southampton Eye Unit, University Hospital Southampton NHS Foundation Trust, Southampton, UK

[11] Manchester Centre for Genomic Medicine, Saint Mary's Hospital, Manchester University NHS Foundation Trust, Manchester, UK

[12] Ophthalmology and Visual Sciences, Medical College of Wisconsin, Milwaukee, WI, USA

.

### SUPPLEMENTARY FIGURES

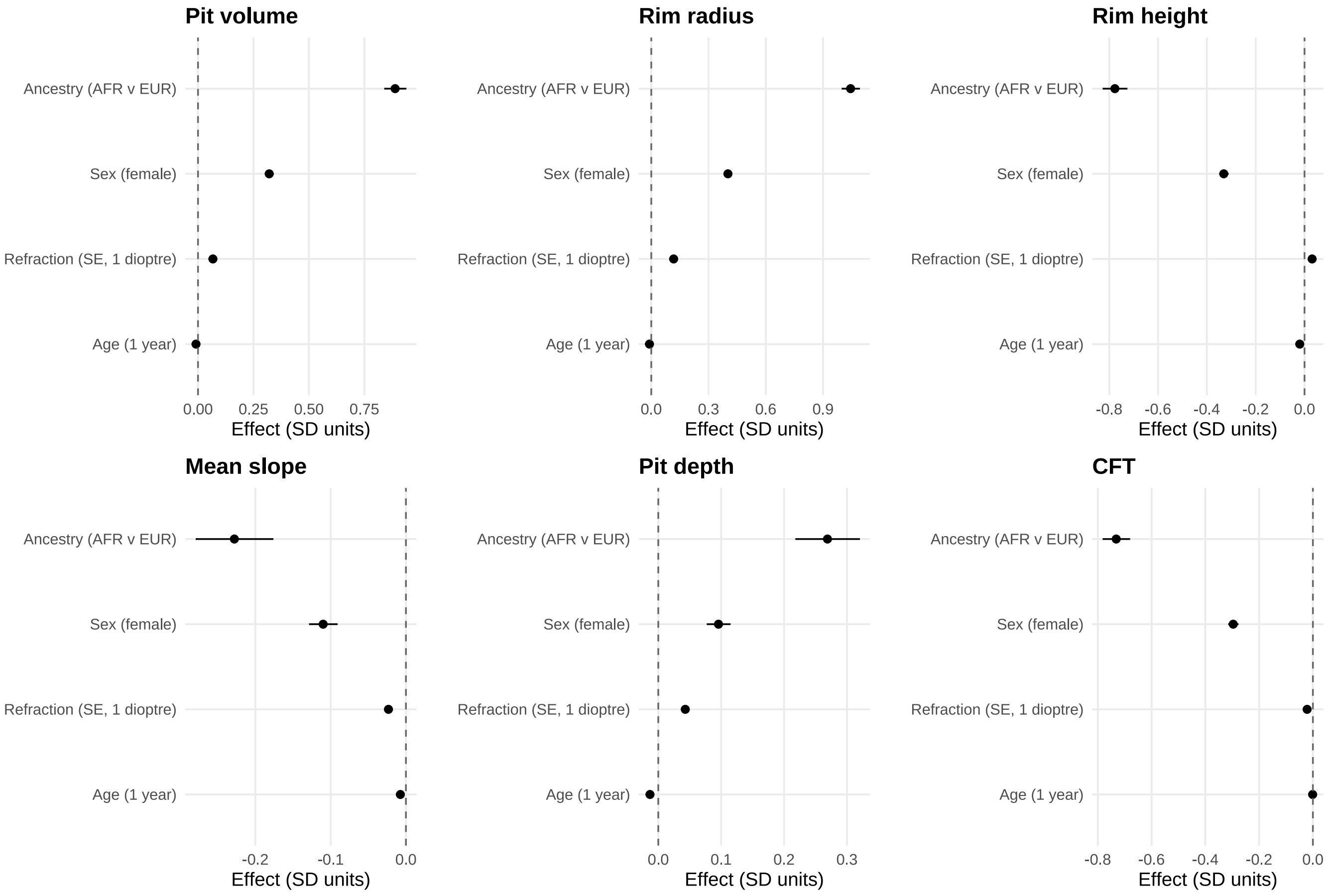


**Supplementary Fig.1.** Forest plots showing the results of multivariate linear regression analyses for foveal traits and demographic features from left and right eye averaged values. Each panel shows the effect estimates from linear regression models fitted separately for the six foveal traits. Predictors included age at scan, spherical equivalent refractive error, sex, and genetic ancestry (African vs. European ancestry compared). Points represent regression coefficients, with horizontal bars indicating 95% confidence intervals. Sex and ancestry showed the largest and most consistent effects across traits, while age and spherical equivalent exhibited comparatively smaller influences. It is noted that averaging was performed by combining left- and right-eye measurements for each trait. When both eyes were available, the mean of the two values was used; when only one eye passed quality control (QC) or was present, that single-eye value was retained.

52

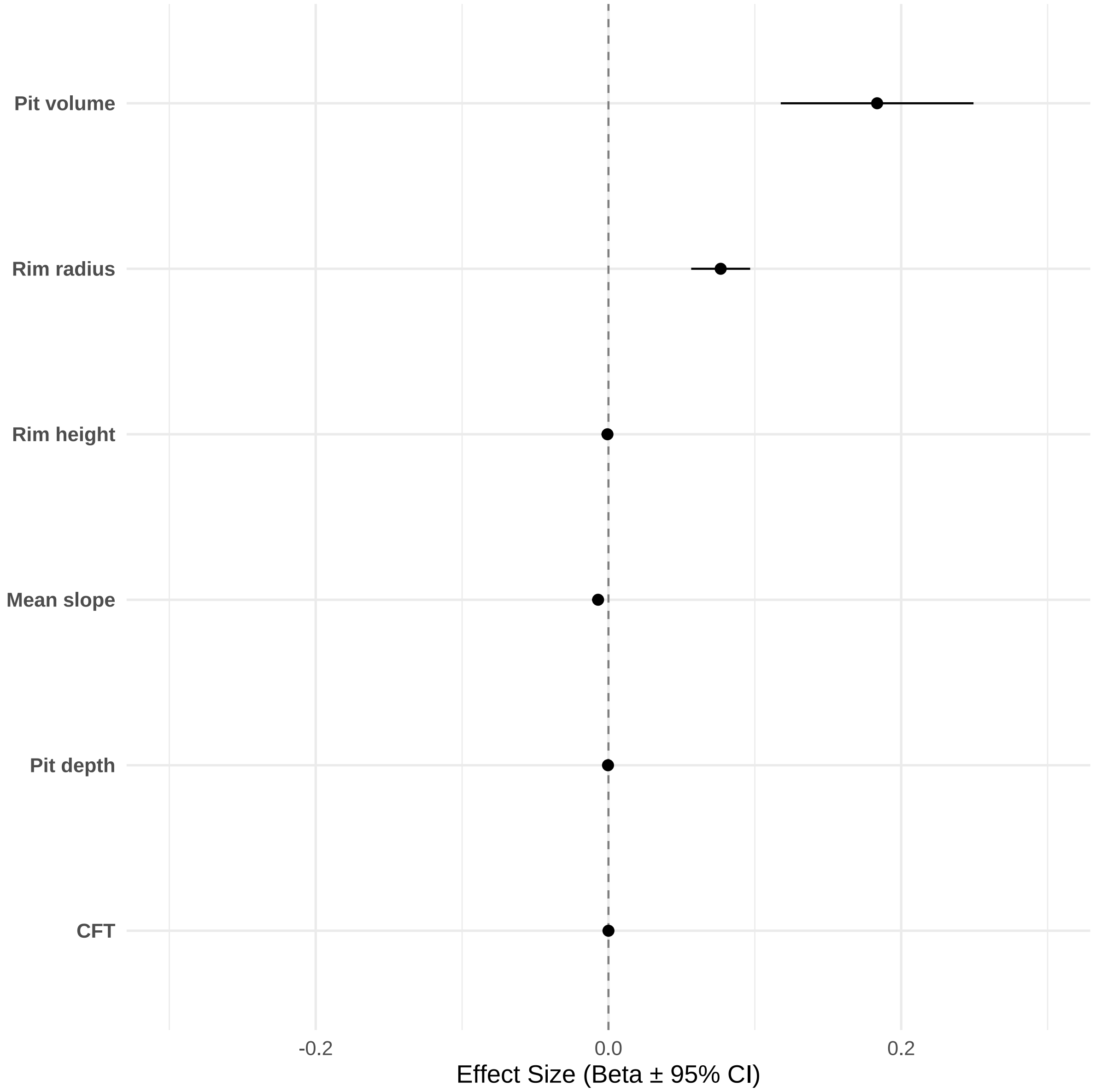


**Supplementary Fig.2.** Association between foveal morphology and visual acuity. Each point shows the beta (β) coefficient (±95% CI) from a linear regression model assessing the effect of a foveal trait on visual acuity (LogMAR), adjusted for age, sex, spherical equivalent refractive error, and genetic ancestry. Negative β values indicate traits linked to better acuity. Rim height ($\beta = -6.8\times10^{-4}$, $p = 1.0\times10^{-23}$, $R^2 = 0.030$) and mean slope ($\beta = -7.1\times10^{-3}$, $p = 4.9\times10^{-21}$, $R^2 = 0.030$) were the strongest predictors, followed by pit depth ($\beta = -3.1\times10^{-4}$, $p = 4.6\times10^{-13}$). Rim radius showed a positive association ($\beta = 7.7\times10^{-2}$, $p = 8.7\times10^{-14}$), indicating that larger radii are linked to worse acuity. Greater pit volume was also associated with worse acuity ($\beta = 0.18$, $p = 4.7\times10^{-8}$). Central foveal thickness (CFT) showed no meaningful effect ($p = 0.92$). Overall, all models explained ~3% of variance, underscoring that foveal morphology accounts for only a small proportion of visual acuity differences in the general population.

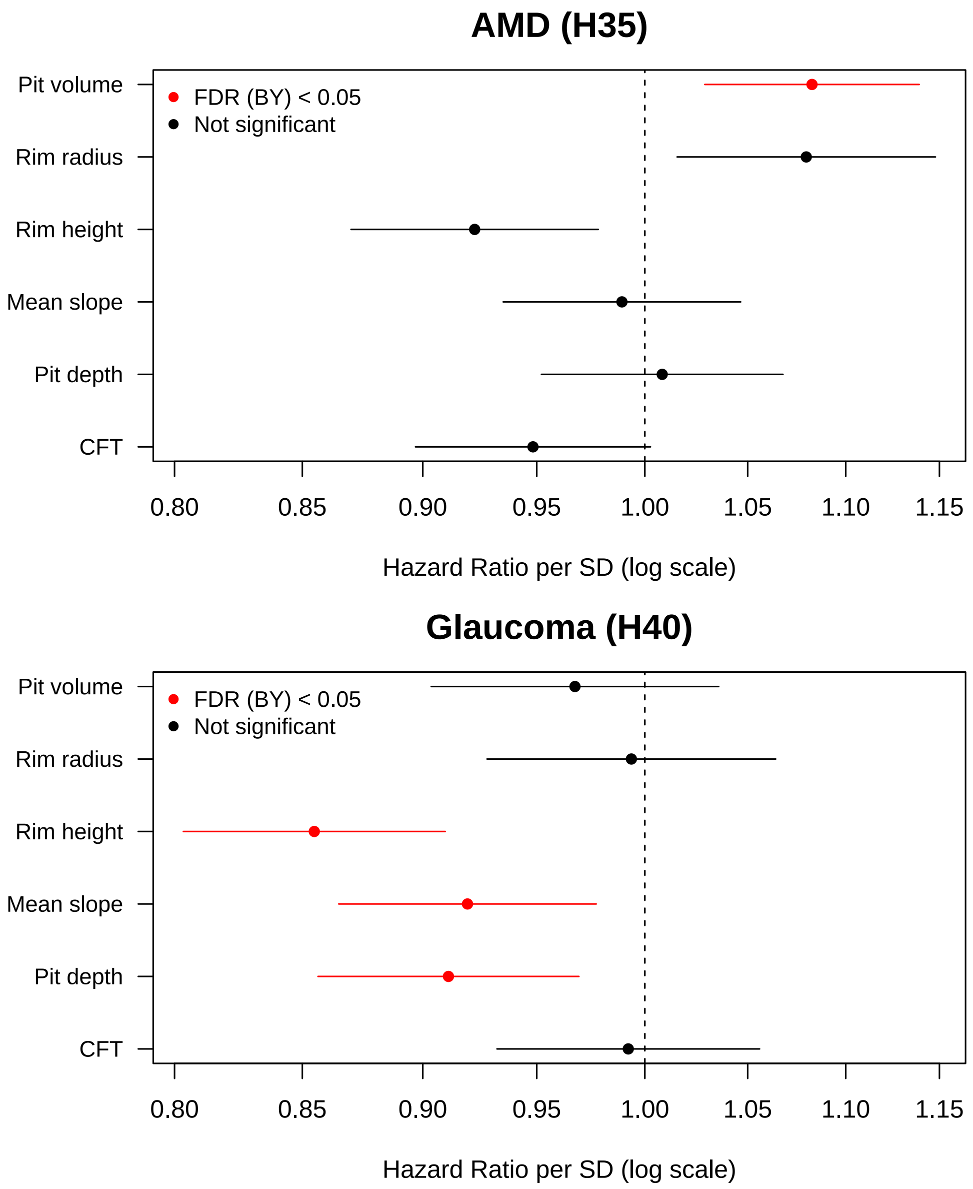


**Supplementary Fig.3.** Cox regression analysis of the associations between averaged foveal traits and post-OCT-scan incident diagnoses of AMD (H35) and glaucoma (H40). Statistically significant associations (after correcting for multiple comparisons using a 5% Benjamini-Yekutieli FDR procedure) are shown in red.

It is noted that averaging was performed by combining left- and right-eye measurements for each trait. When both eyes were available, the mean of the two values was used; when only one eye passed quality control (QC) or was present, that single-eye value was retained.

AMD, age-related macular degeneration; CFT, central foveal thickness; SD, standard deviation; OCT, optical coherence tomography

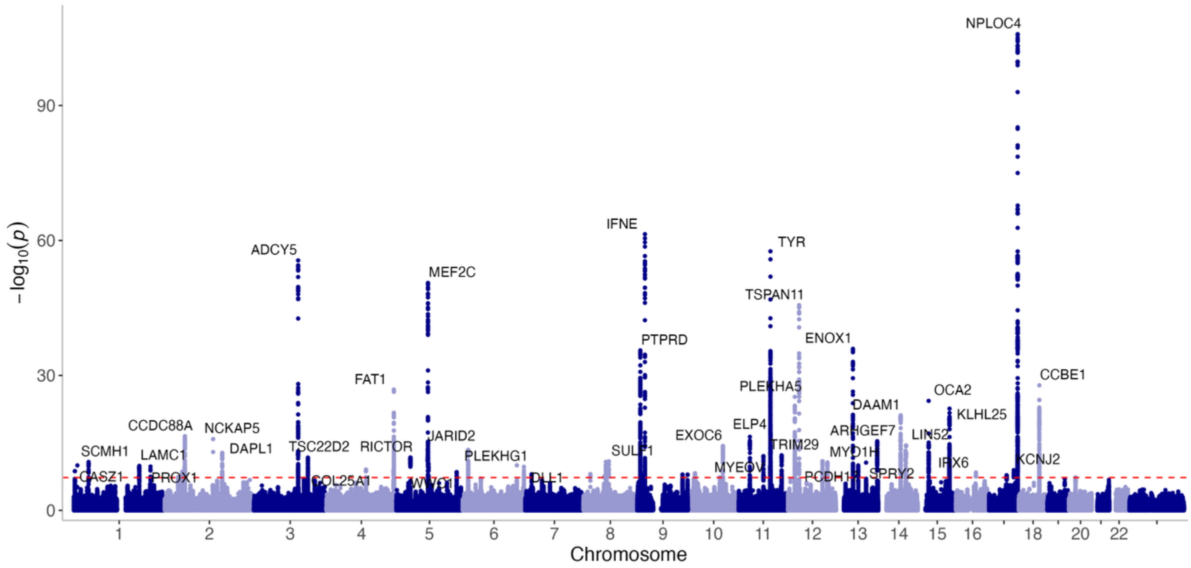


**Supplementary Fig.4.** Manhattan plot showing the findings of a common-variant genome-wide association study (GWAS) for foveal <u>pit volume</u> (left eye). The red line indicates the threshold for genome-wide significance and key lead variants exceeding this value have been annotated.

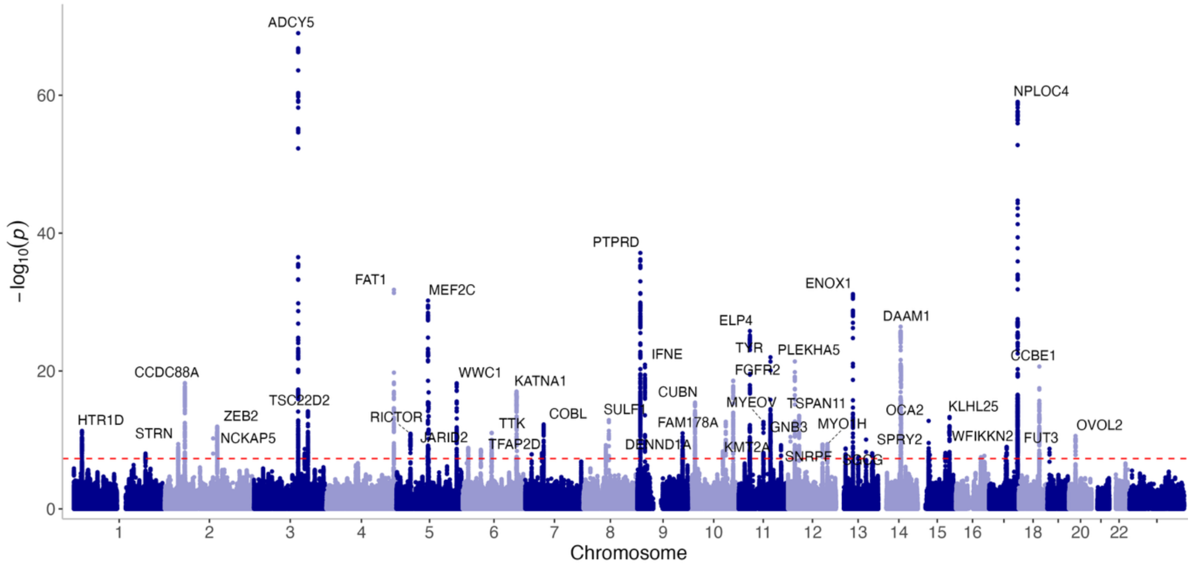


**Supplementary Fig.5.** Manhattan plot showing the findings of a common-variant GWAS for foveal <u>rim radius</u> (left eye).

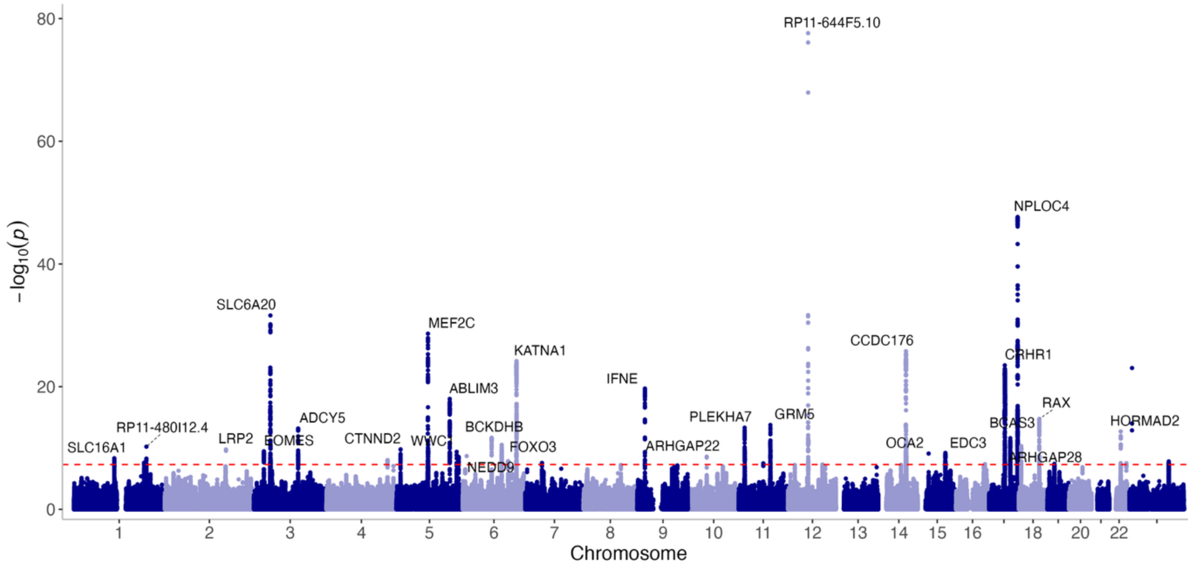


**Supplementary Fig.6.** Manhattan plot showing the findings of a common-variant GWAS for foveal rim height (left eye).

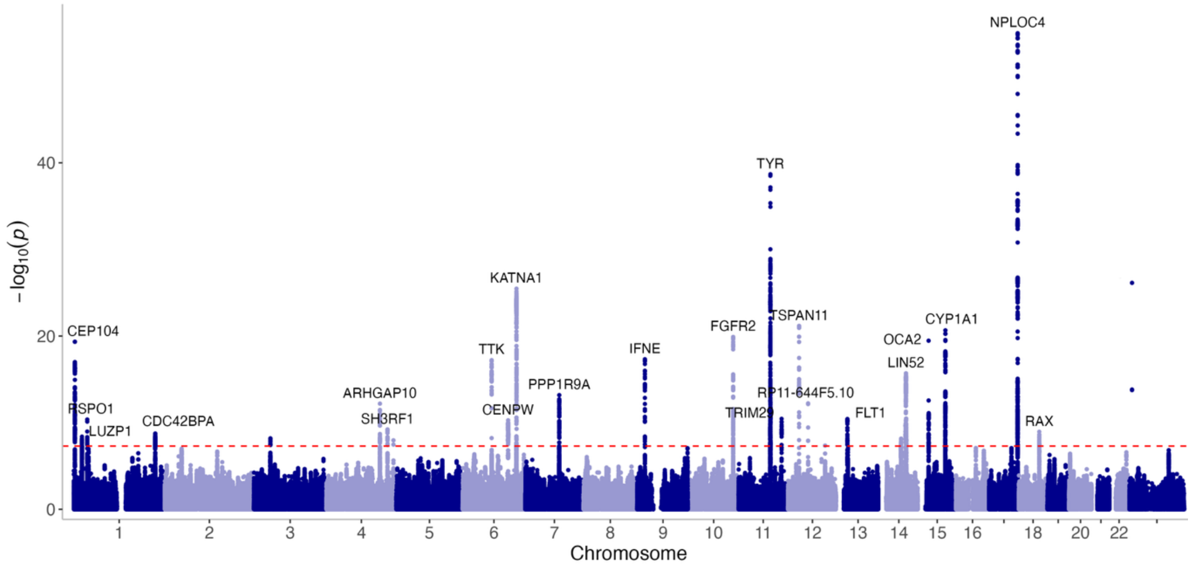


**Supplementary Fig.7.** Manhattan plot showing the findings of a common-variant GWAS for foveal mean slope (left eye).

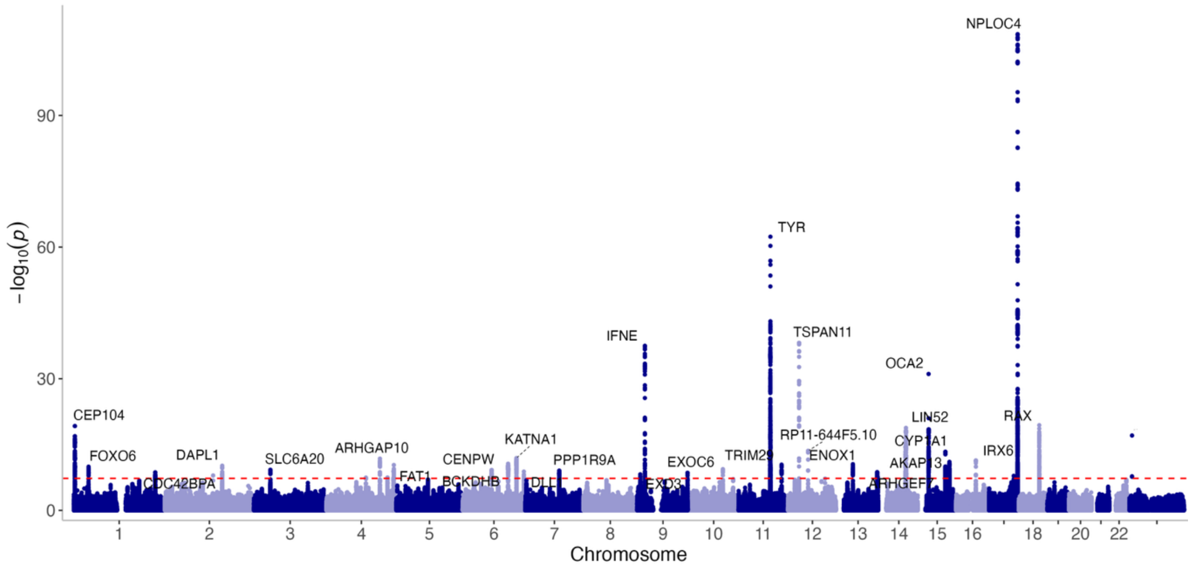


**Supplementary Fig.8.** Manhattan plot showing the findings of a common-variant GWAS for foveal pit depth (left eye).

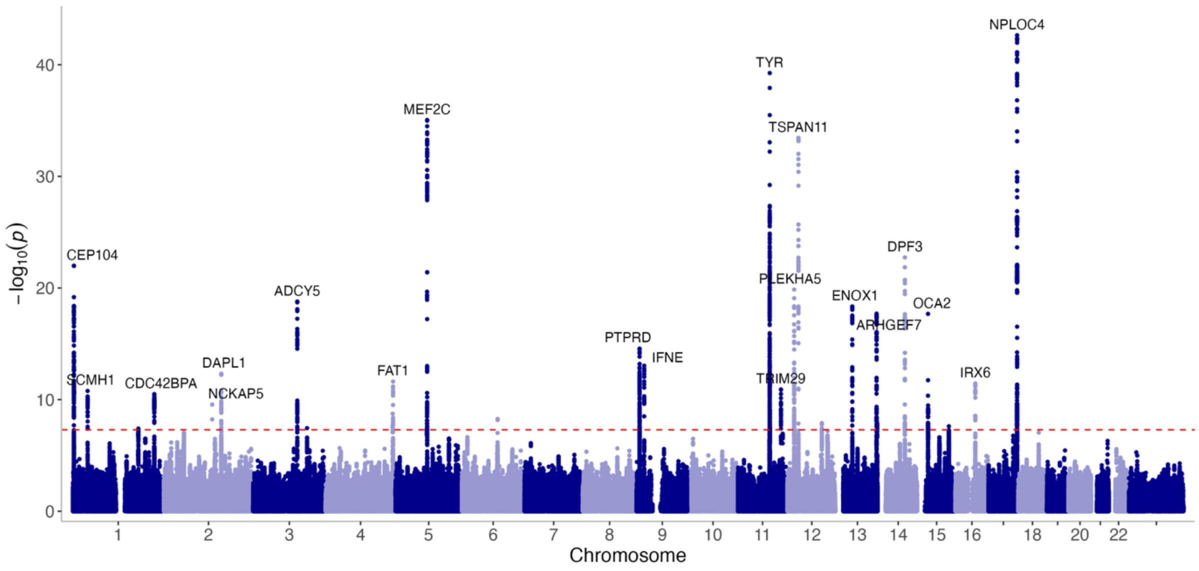


**Supplementary Fig.9.** Manhattan plot showing the findings of a common-variant GWAS for central foveal thickness (CFT) (left eye).

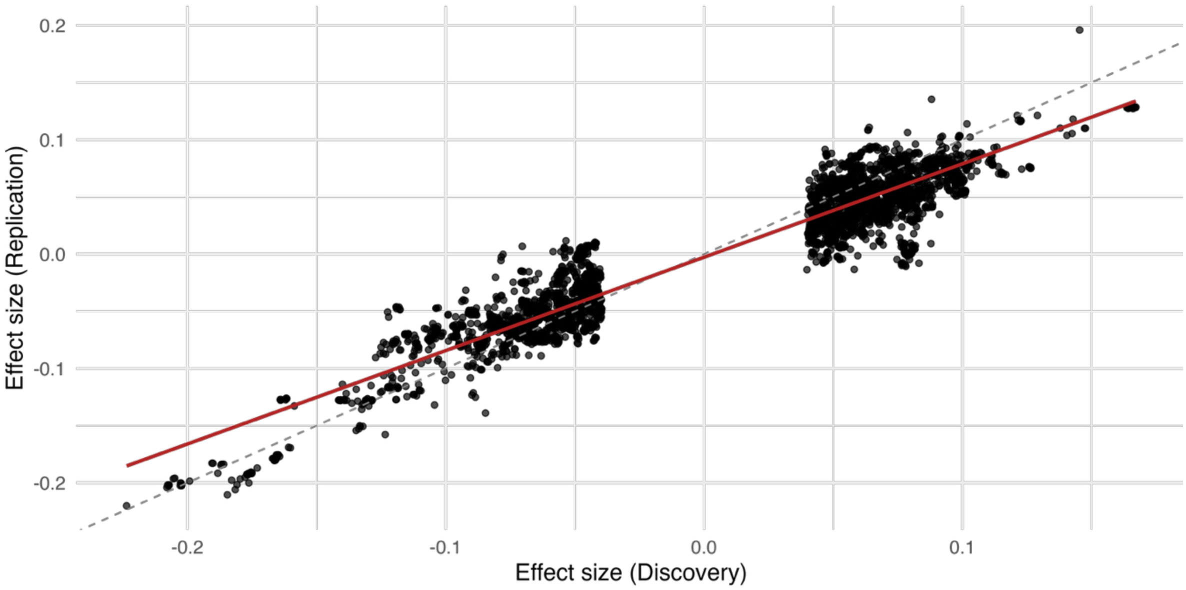


**Supplementary Fig.10.** Beta–beta plot comparing effect size estimates (β) from the primary (n=29,710) and internal validation (n=6,495) cohorts for genome-wide significant variants associated with foveal pit volume (left eye). The dashed line indicates the identity line (y = x), and the red line shows the linear regression fit. The relevant correlation coefficient is $R^2 = 0.94$ ($p < 2.2 \times 10^{-16}$).

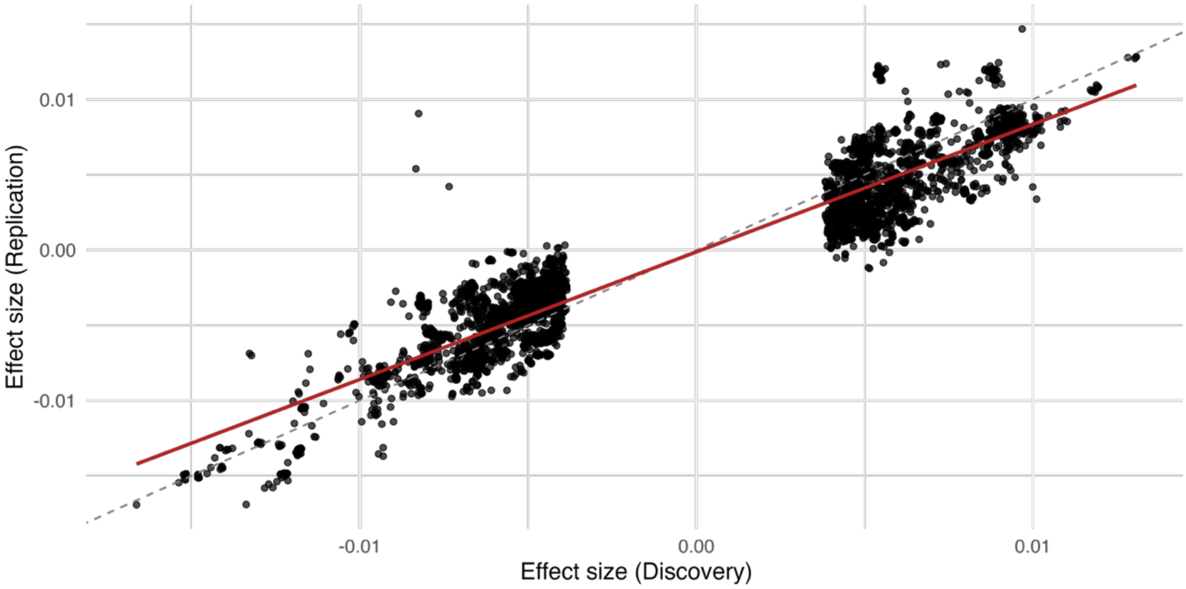


**Supplementary Fig.11.** Beta–beta plot comparing effect size estimates (β) from the discovery and internal validation cohorts for genome-wide significant variants associated with foveal rim radius (left eye). The relevant correlation coefficient is $R^2 = 0.94$ ($p < 2.2 \times 10^{-16}$).

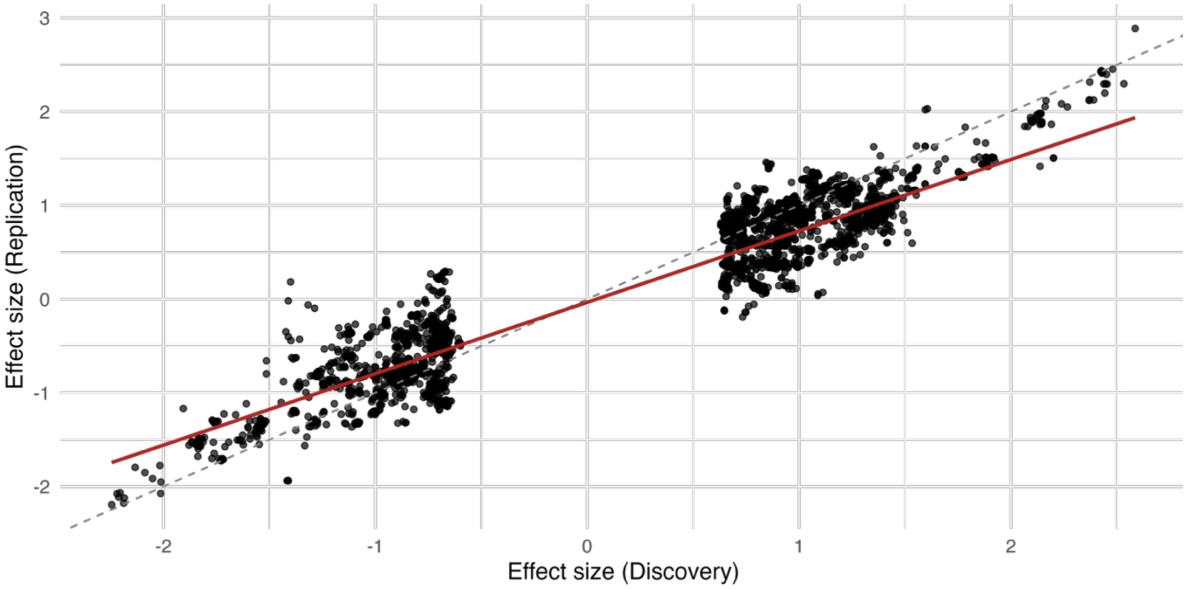


**Supplementary Fig.12.** Beta–beta plot comparing effect size estimates (β) from the discovery and internal validation cohorts for genome-wide significant variants associated with foveal <u>rim height</u> (left eye). The relevant correlation coefficient is $R^2 = 0.95$ ($p < 2.2 \times 10^{-16}$).

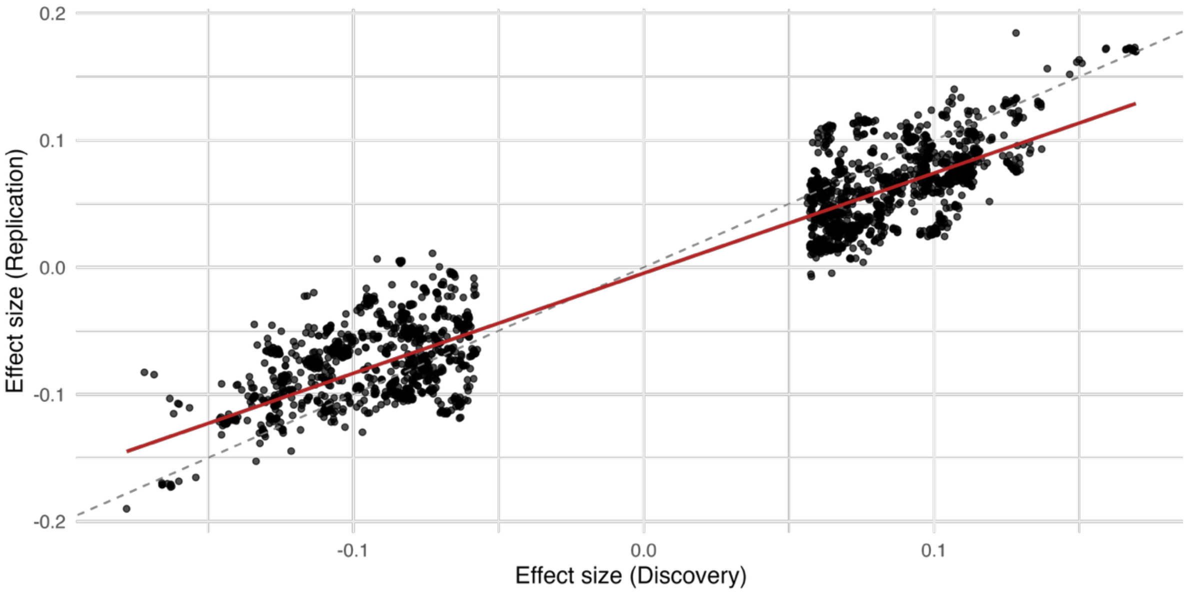


**Supplementary Fig.13.** Beta–beta plot comparing effect size estimates (β) from the discovery and internal validation cohorts for genome-wide significant variants associated with foveal <u>mean slope</u> (left eye). The relevant correlation coefficient is $R^2 = 0.95$ ($p < 2.2 \times 10^{-16}$).

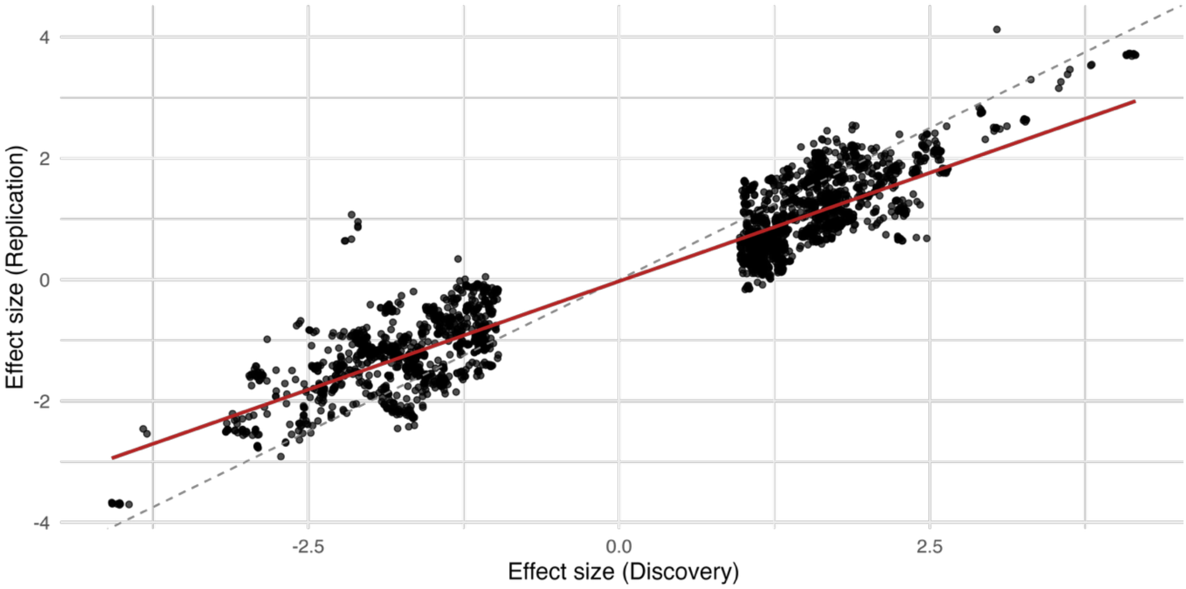


**Supplementary Fig.14.** Beta–beta plot comparing effect size estimates (β) from the discovery and internal validation cohorts for genome-wide significant variants associated with foveal pit depth (left eye). The relevant correlation coefficient is $R^2 = 0.93$ ($p < 2.2 \times 10^{-16}$).

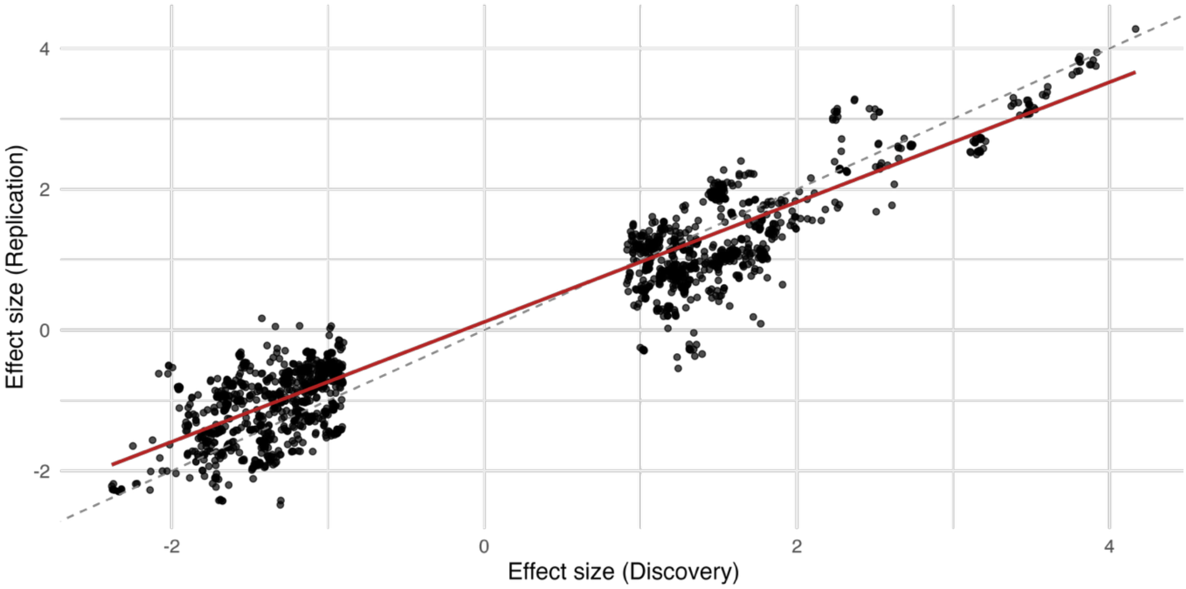


**Supplementary Fig.15.** Beta–beta plot comparing effect size estimates (β) from the discovery and internal validation cohorts for genome-wide significant variants associated with central foveal thickness (CFT) (left eye). Pearson correlation: $R^2 = 0.95$, $p < 2.2 \times 10^{-16}$.

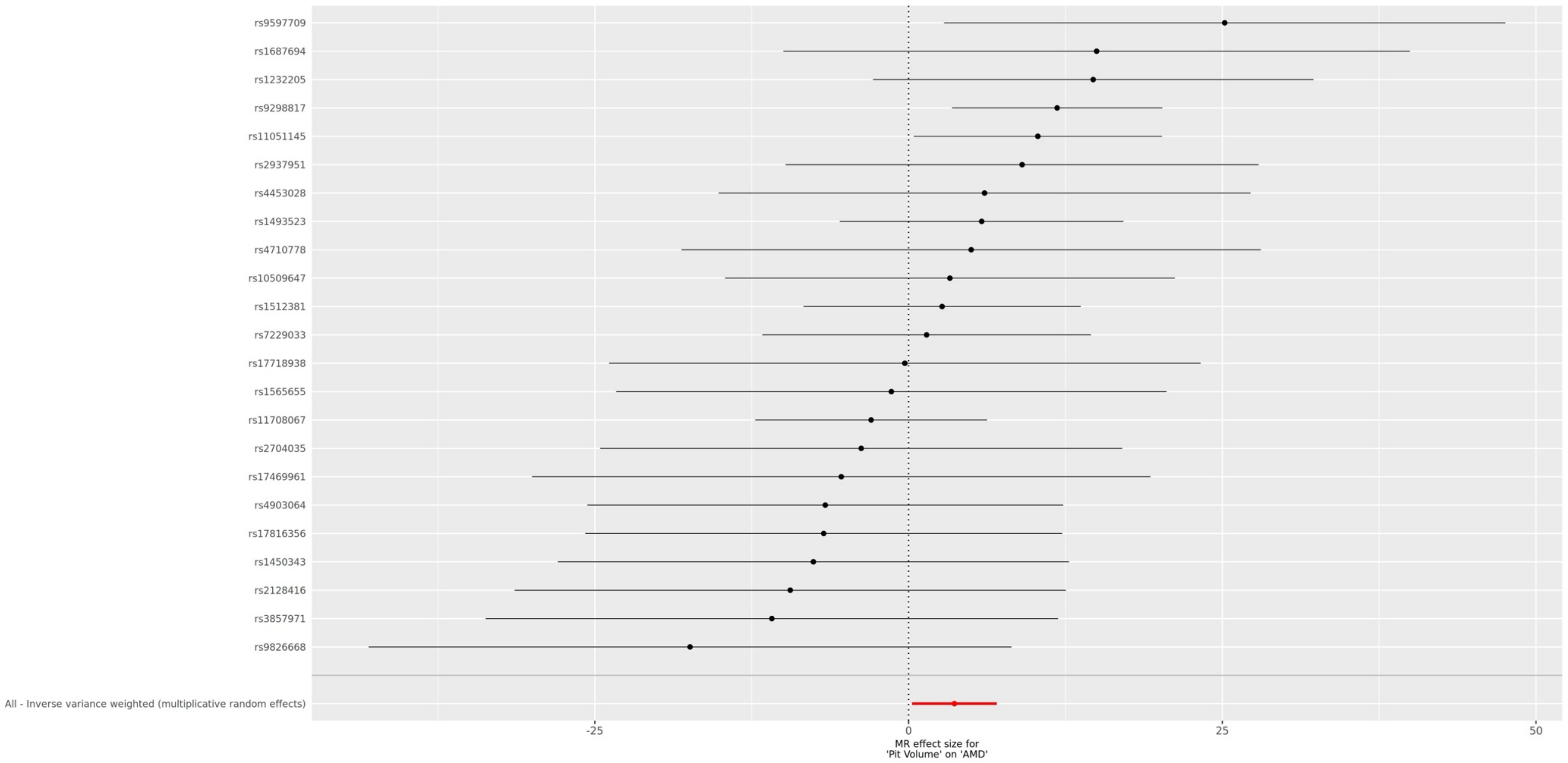


**Supplementary Fig.16:** Forest plot showing variant-level causal estimates for pit volume on age-related macular degeneration (AMD) risk. The overall IVW (inverse-variance weighted) estimate indicates a significant positive association.

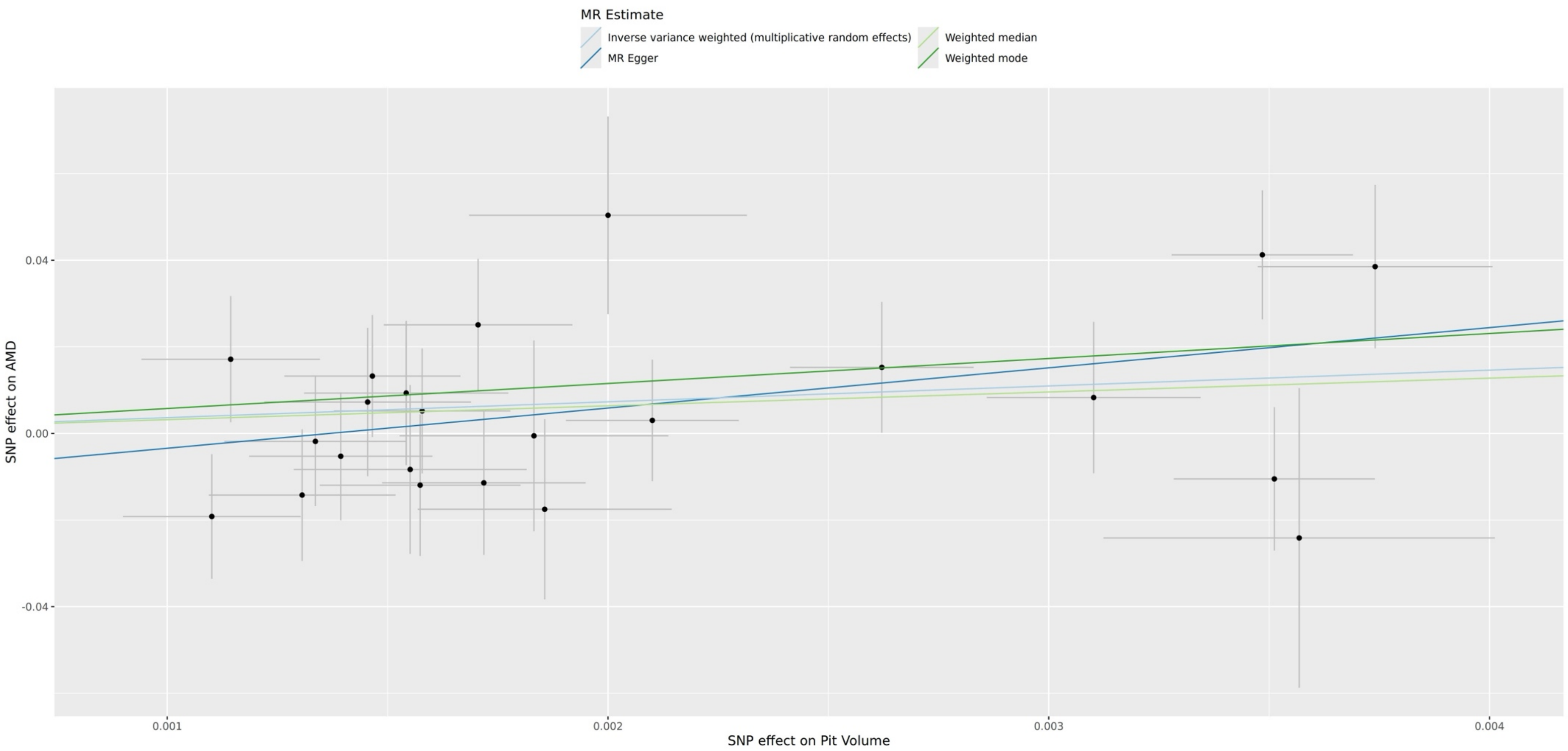


**Supplementary Fig.17:** Mendelian randomization scatter plot for pit volume and age-related macular degeneration (AMD). Points represent variant-specific causal estimates; fitted lines correspond to the IVW (inverse-variance weighted), MR-Egger, weighted median, and weighted mode models.

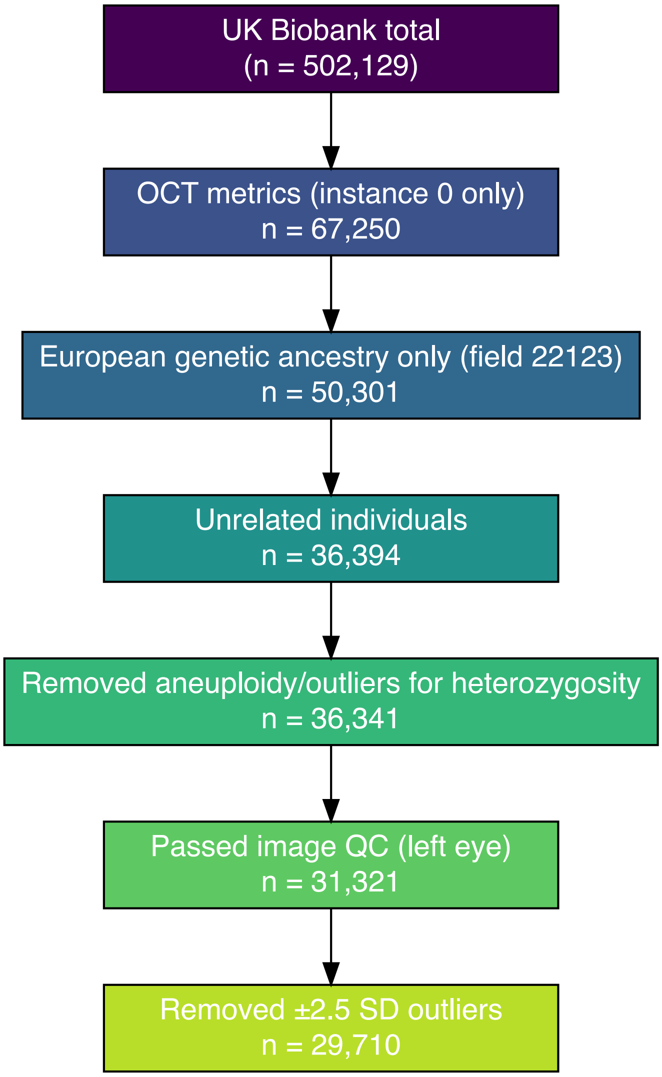


**Supplementary Fig.18:** Overview of the filtering steps used in the primary common-variant GWAS. An identical approach utilizing ‘instance 1’ OCTs was used in the internal validation study**.**

OCT, optical coherence tomography; QC quality control; SD, standard deviation.

### SUPPLEMENTARY TABLES

**Supplementary Table 1.** Multivariate associations between age, refractive error, sex, ancestry, and foveal morphological traits

| **Trait** (left eye) | **Term** | **Estimate** | **Standard Error** | **Statistic** | **P-value** | **Lower 95% CI** | **Upper 95% CI** |
|---|---|---|---|---|---|---|---|
| CFT | Age at scan | 0.0004638 | 0.0006302 | 0.736 | $4.62 \times 10^{-1}$ | -0.0007715 | 0.001699 |
| CFT | Spherical Equivalent | -0.02172 | 0.001947 | -11.156 | $7.40 \times 10^{-29}$ | -0.02553 | -0.01790 |
| CFT | Male sex | 0.2961 | 0.009977 | 29.683 | $1.90 \times 10^{-191}$ | 0.2766 | 0.3157 |
| CFT | Ancestry (AFR) | -0.8026 | 0.02720 | -29.505 | $3.25 \times 10^{-189}$ | -0.8559 | -0.7493 |
| CFT | Ancestry (EAS) | -0.2702 | 0.05021 | -5.382 | $7.43 \times 10^{-8}$ | -0.3686 | -0.1718 |
| CFT | Ancestry (SAS) | -0.5007 | 0.03599 | -13.914 | $6.63 \times 10^{-44}$ | -0.5712 | -0.4302 |
| Pit volume | Age at scan | -0.01211 | 0.0006058 | -19.983 | $2.20 \times 10^{-88}$ | -0.01329 | -0.01092 |
| Pit volume | Spherical Equivalent | 0.07823 | 0.001867 | 41.906 | $p < 1 \times 10^{-300}$ | 0.07457 | 0.08189 |
| Pit volume | Male sex | -0.3435 | 0.009590 | -35.824 | $1.80 \times 10^{-276}$ | -0.3623 | -0.3247 |
| Pit volume | Ancestry (AFR) | 1.008 | 0.02615 | 38.546 | $p < 1 \times 10^{-300}$ | 0.9566 | 1.059 |
| Pit volume | Ancestry (EAS) | 0.6519 | 0.04820 | 13.526 | $1.37 \times 10^{-41}$ | 0.5574 | 0.7464 |
| Pit volume | Ancestry (SAS) | 0.5695 | 0.03466 | 16.434 | $1.77 \times 10^{-60}$ | 0.5016 | 0.6375 |
| Pit depth | Age at scan | -0.01463 | 0.0006380 | -22.934 | $1.30 \times 10^{-115}$ | -0.01588 | -0.01338 |
| Pit depth | Spherical Equivalent | 0.04570 | 0.001974 | 23.156 | $8.22 \times 10^{-118}$ | 0.04183 | 0.04957 |
| Pit depth | Male sex | -0.08347 | 0.01010 | -8.261 | $1.49 \times 10^{-16}$ | -0.1033 | -0.06366 |
| Pit depth | Ancestry (AFR) | 0.2758 | 0.02765 | 9.977 | $2.06 \times 10^{-23}$ | 0.2216 | 0.3300 |
| Pit depth | Ancestry (EAS) | 0.1441 | 0.05105 | 2.823 | $4.76 \times 10^{-3}$ | 0.04405 | 0.2442 |
| Pit depth | Ancestry (SAS) | 0.1090 | 0.03657 | 2.980 | $2.88 \times 10^{-3}$ | 0.03730 | 0.1806 |
| Mean slope | Age at scan | -0.008538 | 0.0006435 | -13.267 | $4.40 \times 10^{-40}$ | -0.009799 | -0.007277 |
| Mean slope | Spherical Equivalent | -0.02147 | 0.001992 | -10.780 | $4.66 \times 10^{-27}$ | -0.02538 | -0.01757 |

| | | | | | | | |
|---|---|---|---|---|---|---|---|
| Mean slope | Male sex | 0.1212 | 0.01019 | 11.896 | $1.42 \times 10^{-32}$ | 0.1012 | 0.1412 |
| Mean slope | Ancestry (AFR) | -0.2146 | 0.02789 | -7.695 | $1.45 \times 10^{-14}$ | -0.2693 | -0.1600 |
| Mean slope | Ancestry (EAS) | -0.1537 | 0.05154 | -2.983 | $2.86 \times 10^{-3}$ | -0.2547 | -0.05270 |
| Mean slope | Ancestry (SAS) | -0.2038 | 0.03694 | -5.518 | $3.45 \times 10^{-8}$ | -0.2763 | -0.1314 |
| Rim radius | Age at scan | -0.01035 | 0.0005820 | -17.780 | $1.94 \times 10^{-70}$ | -0.01149 | -0.009207 |
| Rim radius | Spherical Equivalent | 0.1232 | 0.001798 | 68.519 | $p < 1 \times 10^{-300}$ | 0.1197 | 0.1267 |
| Rim radius | Male sex | -0.4024 | 0.009215 | -43.672 | $p < 1 \times 10^{-300}$ | -0.4205 | -0.3844 |
| Rim radius | Ancestry (AFR) | 1.030 | 0.02519 | 40.882 | $p < 1 \times 10^{-300}$ | 0.9803 | 1.079 |
| Rim radius | Ancestry (EAS) | 0.6745 | 0.04631 | 14.566 | $6.20 \times 10^{-48}$ | 0.5838 | 0.7653 |
| Rim radius | Ancestry (SAS) | 0.6472 | 0.03334 | 19.409 | $1.64 \times 10^{-83}$ | 0.5818 | 0.7125 |
| Rim height | Age at scan | -0.01969 | 0.0006231 | -31.606 | $1.87 \times 10^{-216}$ | -0.02091 | -0.01847 |
| Rim height | Spherical Equivalent | 0.03451 | 0.001910 | 18.068 | $1.14 \times 10^{-72}$ | 0.03077 | 0.03825 |
| Rim height | Male sex | 0.3358 | 0.009861 | 34.053 | $2.06 \times 10^{-250}$ | 0.3165 | 0.3551 |
| Rim height | Ancestry (AFR) | -0.7856 | 0.02707 | -29.024 | $3.24 \times 10^{-183}$ | -0.8387 | -0.7326 |
| Rim height | Ancestry (EAS) | -0.2015 | 0.04942 | -4.078 | $4.55 \times 10^{-5}$ | -0.2984 | -0.1047 |
| Rim height | Ancestry (SAS) | -0.5591 | 0.03575 | -15.640 | $5.73 \times 10^{-55}$ | -0.6292 | -0.4891 |

For each foveal trait, a separate two-sided linear regression model was fitted. Each trait was standardised (Z-scored) prior to modelling, so regression coefficients represent the change in the trait in standard deviation (SD) units per one-unit change in the predictor: per year of age at OCT acquisition, per dioptre of spherical equivalent refractive error, for male relative to female sex, and for each genetic ancestry group relative to European ancestry (the reference level). Positive coefficients indicate higher foveal trait values with increasing predictor values; negative coefficients indicate the opposite. Estimates, standard errors and confidence interval bounds are given to four significant figures. P-values are reported without adjustment for multiple testing. Extremely small p-values that underflow to zero are reported as $p < 1 \times 10^{-300}$. While the contribution of environmental factors cannot be excluded, we focused on established demographic and ocular confounders, as these are known to influence retinal structure and are routinely included in quantitative retinal morphology studies.

**Supplementary Table 2.** Differences in foveal traits between male and female UK Biobank participants

| **Foveal trait** (left eye) | **Male** (n=20,474) | | **Female** (n=19,047) | | **p-value** |
|---|---|---|---|---|---|
| | **mean** | **median** | **mean** | **median** | |
| Pit volume (mm$^3$) | 0.074 | 0.069 | 0.086 | 0.082 | * |
| Rim radius (mm) | 0.954 | 0.946 | 1.001 | 0.996 | * |
| Rim height (µm) | 355.417 | 356.068 | 350.325 | 350.705 | * |
| Mean slope (°) | 6.7 | 6.769 | 6.542 | 6.574 | * |
| Pit depth (µm) | 111.457 | 113.244 | 113.838 | 115.85 | * |
| CFT (µm) | 243.527 | 241.684 | 236.322 | 234.573 | * |

Two-sided tests were used throughout. P-values are reported without correction for multiple testing. A pictorial representation of these left eye results can be found in Fig.3.

* All Wilcoxon rank-sum test p-values were $< 1 \times 10^{-10}$.

**Supplementary Table 3.** Differences in foveal traits between broad genetic ancestry groups

| **Foveal trait** (left eye) | **European** (n=36,972) | | **African** (n=1,386) | | **East Asian** (n=397) | | **South Asian** (n=766) | | **p-value** |
|---|---|---|---|---|---|---|---|---|---|
| | **mean** | **median** | **mean** | **median** | **mean** | **median** | **mean** | **median** | |
| Pit volume ($mm^3$) | 0.078 | 0.074 | 0.114 | 0.11 | 0.1 | 0.093 | 0.097 | 0.093 | * |
| Rim radius (mm) | 0.972 | 0.967 | 1.095 | 1.093 | 1.043 | 1.035 | 1.043 | 1.035 | * |
| Rim height (μm) | 353.34 | 353.674 | 342.295 | 343.078 | 349.93 | 350.097 | 345.735 | 346.205 | * |
| Mean slope (°) | 6.632 | 6.68 | 6.394 | 6.421 | 6.493 | 6.541 | 6.397 | 6.42 | * |
| Pit depth (μm) | 112.281 | 114.289 | 120.896 | 123.418 | 116.445 | 119.809 | 115.659 | 116.878 | * |
| CFT (μm) | 240.775 | 239.117 | 221.111 | 220.5 | 233.581 | 229.994 | 229.398 | 227.562 | * |

A pictorial representation of these left eye results can be found in Fig.4.

* All Kruskal–Wallis p-values were $< 1 \times 10^{-10}$

**Supplementary Table 4.** Changes in adjusted R² associated with inclusion of genetic ancestry and retinal pigment score in models of foveal morphology

| **Foveal trait** (left eye) | **$R^2$** (RPS only) | **p-value** (RPS only) | **$R^2$** (Ancestry only) | **p-value** (Ancestry only) | **$R^2$** (combined) | **p-value** (combined) | **Δ adjusted $R^2$** (adding RPS) | **Δ adjusted $R^2$** (adding ancestry) |
|---|---|---|---|---|---|---|---|---|
| Pit volume | 0.099 | 0 | 0.114 | 0 | 0.119 | 0 | 0.005 | 0.019 |
| Rim radius | 0.153 | 0 | 0.184 | 0 | 0.176 | 0 | -0.008 | 0.023 |
| Rim height | 0.052 | $5.80 \times 10^{-258}$ | 0.069 | 0 | 0.072 | 0 | 0.003 | 0.020 |
| Mean slope | 0.016 | $5.44 \times 10^{-77}$ | 0.013 | $3.30 \times 10^{-151}$ | 0.019 | $6.48 \times 10^{-88}$ | 0.006 | 0.003 |
| Pit depth | 0.039 | $2.21 \times 10^{-190}$ | 0.029 | 0 | 0.039 | $3.91 \times 10^{-190}$ | 0.010 | 0.0005 |
| CFT | 0.038 | $2.23 \times 10^{-187}$ | 0.047 | 0 | 0.051 | $4.11 \times 10^{-251}$ | 0.004 | 0.013 |

Adjusted R² values are shown for models containing RPS alone, ancestry alone, or both predictors. The final two columns show the change in adjusted R² obtained by adding RPS or ancestry to the corresponding nested model. These values are presented as descriptive measures of change in model fit and should not be interpreted as formal unique variance components. Two-sided tests were used throughout. P-values are reported without correction for multiple testing. A pictorial representation of these left eye results can be found in Fig.4.

RPS, retinal pigment score.

**Supplementary Table 5.** Univariable Cox regression analysis results for associations between foveal traits and post-OCT scan incidence of AMD and glaucoma

| **Foveal trait** (left eye) | **Event** | **Number of cases** | **Hazard ratio (per SD)** | **Lower 95% CI (per SD)** | **Upper 95% CI (per SD)** | **p-value** | **p-value (FDR)** |
|---|---|---|---|---|---|---|---|
| Pit volume | AMD | 956 | 1.097 | 1.035 | 1.163 | 0.002 | 0.026 |
| Rim radius | AMD | 953 | 1.055 | 0.986 | 1.129 | 0.123 | 0.451 |
| Rim height | AMD | 959 | 0.932 | 0.874 | 0.994 | 0.033 | 0.239 |
| Mean slope | AMD | 962 | 0.967 | 0.909 | 1.029 | 0.288 | 0.848 |
| Pit depth | AMD | 958 | 1.005 | 0.944 | 1.071 | 0.865 | 1 |
| CFT | AMD | 949 | 0.945 | 0.888 | 1.006 | 0.076 | 0.374 |
| Pit volume | Glaucoma | 841 | 0.927 | 0.860 | 1.000 | 0.050 | 0.185 |
| Rim radius | Glaucoma | 836 | 0.976 | 0.906 | 1.052 | 0.525 | 1 |
| Rim height | Glaucoma | 844 | 0.830 | 0.776 | 0.888 | $5.68 \times 10^{-8}$ | $8.35 \times 10^{-7}$ |
| Mean slope | Glaucoma | 836 | 0.894 | 0.838 | 0.955 | $8.76 \times 10^{-4}$ | 0.004 |
| Pit depth | Glaucoma | 837 | 0.874 | 0.818 | 0.933 | $6.16 \times 10^{-5}$ | $4.53 \times 10^{-4}$ |
| CFT | Glaucoma | 839 | 1.008 | 0.942 | 1.078 | 0.823 | 1 |

Two-sided regression analyses were performed in a cohort of 39,521 individuals, and all models were adjusted for age at image acquisition, sex, spherical equivalent refractive error and genetic ancestry. Hazard ratios and confidence intervals are all expressed per one standard deviation. All numbers are rounded to three decimal places. A pictorial representation of these left eye results can be found in Fig.5.

AMD, age-related macular degeneration; CI, confidence interval; FDR, false discovery rate corrected; SD, standard deviation.

**Supplementary Table 6.** Heritability and genomic inflation estimates for the six studied foveal traits

| **Foveal trait** (left eye) | **$h^2$** | **$h^2$ standard error** | **λGC** | **Intercept** | **Intercept standard error** | **Ratio** | **Ratio standard error** |
|---|---|---|---|---|---|---|---|
| Pit volume | 0.427 | 0.039 | 1.165 | 1.027 | 0.009 | 0.095 | 0.034 |
| Rim radius | 0.390 | 0.034 | 1.159 | 1.027 | 0.009 | 0.100 | 0.037 |
| Rim height | 0.412 | 0.038 | 1.156 | 1.005 | 0.010 | 0.020 | 0.039 |
| Mean slope | 0.311 | 0.036 | 1.120 | 1.014 | 0.009 | 0.065 | 0.047 |
| Pit depth | 0.338 | 0.040 | 1.130 | 1.020 | 0.010 | 0.089 | 0.045 |
| CFT | 0.287 | 0.028 | 1.130 | 1.020 | 0.009 | 0.106 | 0.048 |

SNV-based heritability ($h^2$) estimates were calculated using LD score regression. All traits showed moderate to high heritability, with pit volume and rim height exhibiting the highest values. The LD score intercepts remained close to 1 and attenuation ratios were low (<0.11), indicating that most inflation reflects true polygenic signal rather than confounding.

SNV, single nucleotide variant, LD, linkage disequilibrium.

**Supplementary Table 7.** Genetic correlation studies involving retinal pigment score and the six studied foveal traits

| Trait 1 | Trait 2 | Correlation coefficient (rg) | Standard error | z-score | p-value |
|---|---|---|---|---|---|
| RPS | CFT | 0.228 | 0.092 | 2.49 | 0.013 |
| RPS | Mean slope | -0.221 | 0.087 | -2.543 | 0.011 |
| RPS | Pit depth | -0.234 | 0.097 | -2.4 | 0.016 |
| RPS | Pit volume | -0.118 | 0.085 | -1.387 | 0.166 |
| RPS | Rim height | -0.074 | 0.073 | -1.006 | 0.315 |
| RPS | Rim radius | -0.035 | 0.081 | -0.429 | 0.668 |
| CFT | RPS | 0.228 | 0.092 | 2.49 | 0.013 |
| CFT | Mean slope | -0.474 | 0.054 | -8.718 | $2.83 \times 10^{-18}$ |
| CFT | Pit depth | -0.693 | 0.038 | -18.298 | $8.58 \times 10^{-75}$ |
| CFT | Pit volume | -0.786 | 0.032 | -24.557 | $3.61 \times 10^{-133}$ |
| CFT | Rim height | 0.198 | 0.070 | 2.833 | 0.005 |
| CFT | Rim radius | -0.513 | 0.049 | -10.492 | $9.36 \times 10^{-26}$ |
| Mean slope | RPS | -0.221 | 0.087 | -2.543 | 0.011 |
| Mean slope | CFT | -0.474 | 0.054 | -8.718 | $2.83 \times 10^{-18}$ |
| Mean slope | Pit depth | 0.881 | 0.018 | 50.373 | 0 |
| Mean slope | Pit volume | 0.366 | 0.067 | 5.44 | $5.32 \times 10^{-8}$ |
| Mean slope | Rim height | 0.646 | 0.039 | 16.738 | $6.96 \times 10^{-63}$ |
| Mean slope | Rim radius | -0.153 | 0.077 | -1.978 | 0.048 |
| Pit depth | RPS | -0.234 | 0.097 | -2.4 | 0.016 |
| Pit depth | CFT | -0.693 | 0.038 | -18.298 | $8.58 \times 10^{-75}$ |

(continued)

(continued)

| Trait 1 | Trait 2 | Correlation coefficient (rg) | Standard error | z-score | p-value |
|---|---|---|---|---|---|
| Pit depth | Mean slope | 0.881 | 0.018 | 50.373 | 0 |
| Pit depth | Pit volume | 0.757 | 0.032 | 23.428 | $2.2 \times 10^{-121}$ |
| Pit depth | Rim height | 0.569 | 0.044 | 12.773 | $2.33 \times 10^{-37}$ |
| Pit depth | Rim radius | 0.338 | 0.062 | 5.447 | $5.11 \times 10^{-8}$ |
| Pit volume | RPS | -0.118 | 0.085 | -1.387 | 0.166 |
| Pit volume | CFT | -0.786 | 0.032 | -24.557 | $3.61 \times 10^{-133}$ |
| Pit volume | Mean slope | 0.366 | 0.067 | 5.44 | $5.32 \times 10^{-8}$ |
| Pit volume | Pit depth | 0.757 | 0.032 | 23.428 | $2.2 \times 10^{-121}$ |
| Pit volume | Rim height | 0.137 | 0.063 | 2.185 | 0.029 |
| Pit volume | Rim radius | 0.851 | 0.017 | 49.399 | 0 |
| Rim height | RPS | -0.074 | 0.073 | -1.006 | 0.315 |
| Rim height | CFT | 0.198 | 0.07 | 2.833 | 0.005 |
| Rim height | Mean slope | 0.646 | 0.039 | 16.738 | $6.96 \times 10^{-63}$ |
| Rim height | Pit depth | 0.569 | 0.044 | 12.773 | $2.33 \times 10^{-37}$ |
| Rim height | Pit volume | 0.137 | 0.063 | 2.185 | 0.029 |
| Rim height | Rim radius | -0.115 | 0.06 | -1.94 | 0.052 |
| Rim radius | RPS | -0.035 | 0.081 | -0.429 | 0.668 |
| Rim radius | CFT | -0.513 | 0.049 | -10.492 | $9.36 \times 10^{-26}$ |
| Rim radius | Mean slope | -0.153 | 0.077 | -1.978 | 0.048 |
| Rim radius | Pit depth | 0.338 | 0.062 | 5.447 | $5.11 \times 10^{-8}$ |
| Rim radius | Pit volume | 0.851 | 0.017 | 49.399 | 0 |
| Rim radius | Rim height | -0.115 | 0.06 | -1.94 | 0.052 |

Two-sided tests were used throughout. P-values are reported without correction for multiple testing. A pictorial representation of some of these left eye results can be found in Fig.8.

**Supplementary Table 8:** Two-sample Mendelian randomization (MR) analyses assessing causal relationships between foveal traits and retinal disease.

| Exposure | Outcome | Total number of SNPs | Odds ratio | Lower 95% CI | Upper 95% CI | MRE IVW p-value | Weighted median p-value | Egger p-value | Cochran's Q p-value | Egger intercept p-value | Leave one out |
|---|---|---|---|---|---|---|---|---|---|---|---|
| AMD | Pit volume | 36 | 1.000 | 1.000 | 1.000 | 0.621 | 0.262 | 0.394 | 0.535 | 0.477 | Not robust |
| Pit volume | AMD | 23 | 38.807 | 1.318 | 1142.296 | 0.034 | 0.205 | 0.050 | 0.336 | 0.187 | Not robust |
| AMD | Rim radius | 40 | 1.000 | 0.999 | 1.001 | 0.622 | 0.908 | 0.996 | 0.513 | 0.695 | Not robust |
| Rim radius | AMD | 22 | 1.254 | 0.515 | 3.052 | 0.618 | 0.901 | 0.982 | 0.835 | 0.854 | Not robust |
| AMD | Rim height | 34 | 1.019 | 0.778 | 1.334 | 0.891 | 0.727 | 0.402 | 0.251 | 0.259 | Not robust |
| Rim height | AMD | 13 | 1.010 | 1.003 | 1.017 | 0.003 | 0.166 | 0.115 | 0.864 | 0.334 | All significant |
| Mean slope | POAG | 22 | 0.985 | 0.919 | 1.055 | 0.660 | 0.583 | 0.192 | 0.530 | 0.215 | Not robust |
| POAG | Mean slope | 34 | 1.010 | 0.975 | 1.047 | 0.564 | 0.464 | 0.877 | 0.341 | 0.727 | Not robust |
| Pit depth | POAG | 29 | 0.999 | 0.995 | 1.002 | 0.423 | 0.297 | 0.044 | 0.658 | 0.058 | Not robust |
| POAG | Pit depth | 36 | 0.840 | 0.460 | 1.532 | 0.570 | 0.604 | 0.095 | 0.226 | 0.115 | Not robust |
| Rim height | POAG | 28 | 1.005 | 1.000 | 1.011 | 0.068 | 0.117 | 0.663 | 0.396 | 0.798 | Not robust |
| POAG | Rim height | 36 | 0.766 | 0.561 | 1.046 | 0.093 | 0.530 | 0.532 | 0.692 | 0.944 | Not robust |

Each row represents a single two-sided Mendelian randomisation analysis; the corresponding exposure and outcome are specified in the first two columns. Exposure–outcome pairs were included based on statistically significant associations in univariate Cox models (Supplementary Table 5), with the pit volume–AMD analysis undertaken as a follow-up to the corresponding survival analysis. Both directions of association were tested to ensure that reverse causation was correction interrogated. The number of SNVs used as instruments for the exposure is indicated. The primary analysis was performed using the multiplicative random-effects inverse variance weighted (MRE IVW) method, with p-values reported alongside those from three robust Mendelian randomization methods: weighted median, MR Egger and weighted mode. Evidence of heterogeneity (Cochran's Q p-value) and directional pleiotropy (Egger intercept p-value) are also shown. The final column summarizes whether the MRE IVW was consistently significant throughout a leave-one-out analysis. P-values are reported in their unadjusted form.

All instruments were genome-wide significant ($p < 5 \times 10^{-8}$), LD-clumped ($R^2 < 0.001$, 10 Mb window), harmonized between exposure and outcome datasets, and screened for outliers and pleiotropic variants using radial Mendelian randomization and Steiger filtering respectively. Palindromic SNVs with a minor allele frequency (MAF) > 0.42 were excluded to avoid strand ambiguity. Summary statistics for AMD and POAG were obtained from Han *et al.*[1] and Gharahkhani *et al.*[2], respectively. See Methods for further details.

MTAG, Multi-Trait Analysis of GWAS

**Supplementary Table 9.** Leave-one-out IVW analysis testing for influential SNPs in the pit volume–AMD causal model

| Exposure | Outcome | SNP | b | se | p-value |
|---|---|---|---|---|---|
| Pit Volume | AMD | rs17816356 | 3.97 | 1.75 | 0.02 |
| Pit Volume | AMD | rs2128416 | 3.95 | 1.73 | 0.02 |
| Pit Volume | AMD | rs2704035 | 3.84 | 1.77 | 0.03 |
| Pit Volume | AMD | rs2937951 | 3.50 | 1.78 | 0.05 |
| Pit Volume | AMD | rs3857971 | 3.96 | 1.72 | 0.02 |
| Pit Volume | AMD | rs4453028 | 3.60 | 1.79 | 0.04 |
| Pit Volume | AMD | rs4710778 | 3.63 | 1.78 | 0.04 |
| Pit Volume | AMD | rs4903064 | 3.97 | 1.75 | 0.02 |
| Pit Volume | AMD | rs7229033 | 3.80 | 1.82 | 0.04 |
| Pit Volume | AMD | rs9298817 | 2.24 | 1.74 | 0.20 |
| Pit Volume | AMD | rs9597709 | 3.20 | 1.65 | 0.05 |
| Pit Volume | AMD | rs9826668 | 4.00 | 1.68 | 0.02 |
| Pit Volume | AMD | All | 3.66 | 1.73 | 0.03 |

AMD, age-related macular degeneration.

**Supplementary Table 10.** Logistic regression analysis investigating the association between the six studied foveal traits and prevalent AMD and glaucoma

| Foveal trait (left eye) | Event | Number of cases | Odds ratio (per SD) | Lower 95% CI (per SD) | Upper 95% CI (per SD) | p-value | p-value (FDR) |
|---|---|---|---|---|---|---|---|
| Pit volume | AMD | 1164 | 1.108 | 1.051 | 1.169 | $1.64 \times 10^{-4}$ | 0.002 |
| Rim radius | AMD | 1152 | 1.041 | 0.977 | 1.109 | 0.213 | 0.627 |
| Rim height | AMD | 1162 | 0.943 | 0.889 | 1.001 | 0.054 | 0.267 |
| Mean slope | AMD | 1165 | 1.004 | 0.949 | 1.063 | 0.878 | 1 |
| Pit depth | AMD | 1165 | 1.039 | 0.980 | 1.101 | 0.197 | 0.627 |
| CFT | AMD | 1156 | 0.906 | 0.857 | 0.959 | 0.001 | 0.004 |
| Pit volume | Glaucoma | 1175 | 0.926 | 0.868 | 0.988 | 0.020 | 0.098 |
| Rim radius | Glaucoma | 1168 | 0.954 | 0.894 | 1.018 | 0.154 | 0.360 |
| Rim height | Glaucoma | 1179 | 0.846 | 0.798 | 0.897 | $1.77 \times 10^{-8}$ | $1.04 \times 10^{-7}$ |
| Mean slope | Glaucoma | 1174 | 0.922 | 0.871 | 0.976 | 0.005 | 0.041 |
| Pit depth | Glaucoma | 1176 | 0.892 | 0.842 | 0.944 | $8.09 \times 10^{-5}$ | 0.001 |
| CFT | Glaucoma | 1178 | 1.001 | 0.945 | 1.061 | 0.962 | 1 |

Analyses were performed in a cohort of 39,521 individuals and all models were adjusted for age at image acquisition, sex, spherical equivalent refractive error and genetic ancestry. Odds ratios and confidence intervals are reported per one standard deviation increase in the relevant foveal trait. Two-sided tests were used throughout.

**Supplementary Table 11.** Sensitivity analysis of hazard ratios from Cox regression after exclusion of spherical equivalent outliers

| **Foveal trait** (left eye) | **Disease** | **HR_raw** | **HR_6D** | **HR_2D** | **HR_1D** |
|---|---|---|---|---|---|
| Pit volume | AMD (H35) | 1.097 | 1.109 | 1.105 | 1.095 |
| Rim radius | AMD (H35) | 1.055 | 1.077 | 1.071 | 1.153 |
| Rim height | AMD (H35) | 0.932 | 0.916 | 0.917 | 0.886 |
| Mean slope | AMD (H35) | 0.967 | 0.945 | 0.935 | 0.934 |
| Pit depth | AMD (H35) | 1.005 | 0.991 | 0.992 | 0.987 |
| CFT | AMD (H35) | 0.945 | 0.929 | 0.923 | 0.914 |
| Pit volume | Glaucoma (H40) | 0.927 | 0.925 | 0.943 | 0.986 |
| Rim radius | Glaucoma (H40) | 0.976 | 0.981 | 0.991 | 1.023 |
| Rim height | Glaucoma (H40) | 0.830 | 0.810 | 0.799 | 0.772 |
| Mean slope | Glaucoma (H40) | 0.894 | 0.888 | 0.874 | 0.920 |
| Pit depth | Glaucoma (H40) | 0.874 | 0.868 | 0.846 | 0.917 |
| CFT | Glaucoma (H40) | 1.008 | 0.995 | 0.997 | 0.917 |

AMD, age-related macular degeneration.

**Supplementary Table 12:** Mean and median values of foveal morphological traits for the left eye, right eye and the average measurement

| Foveal trait | Left | | Right | | Average | |
|---|---|---|---|---|---|---|
| | Mean | Median | Mean | Median | Mean | Median |
| **CFT (μm)** | 240 | 238 | 238 | 238 | 239 | 238 |
| **Mean slope (°)** | 6.62 | 6.66 | 6.64 | 6.66 | 6.62 | 6.65 |
| **Pit depth (μm)** | 113 | 115 | 114 | 115 | 113 | 115 |
| **Pit volume ($mm^3$)** | 0.0801 | 0.0754 | 0.0837 | 0.0769 | 0.0823 | 0.0766 |
| **Rim height (μm)** | 353 | 353 | 352 | 353 | 352 | 353 |
| **Rim radius (mm)** | 0.979 | 0.972 | 0.985 | 0.976 | 0.982 | 0.975 |

Although statistical differences between eyes were detected (all $p < 0.05$; two-sided tests; no adjustment for multiple testing), the magnitude of these differences was not considered biologically meaningful.

For averaged analyses, left- and right-eye measurements were combined for each trait. When both eyes passed quality control (QC), the mean of the two values was used; when only one eye passed quality control or was available, the corresponding single-eye measurement was retained.
